# White Blood Cell and Platelet Dynamics Define Human Inflammatory Recovery

**DOI:** 10.1101/2021.06.19.21259181

**Authors:** Brody H Foy, Thor Sundt, Jonathan C T Carlson, Aaron D Aguirre, John M Higgins

## Abstract

Inflammation is the physiologic reaction to cellular and tissue damage caused by pathologic processes including trauma, infection, and ischemia^1^. Effective inflammatory responses integrate molecular and cellular functions to prevent further tissue damage, initiate repair, and restore homeostasis, while futile or dysfunctional responses allow escalating injury, delay recovery, and may hasten death^2^. Elevation of white blood cell count (WBC) and altered levels of other acute phase reactants are cardinal signs of inflammation, but the dynamics of these changes and their resolution are not established^3,4^. Patient responses appear to vary dramatically with no clearly defined signs of good prognosis, leaving physicians reliant on qualitative interpretations of laboratory trends^4,5^.

We retrospectively, observationally studied the human acute inflammatory response to trauma, ischemia, and infection by tracking the longitudinal dynamics of cellular and serum markers in hospitalized patients. Unexpectedly, we identified a conserved pattern of recovery defined by co-regulation of WBC and platelet (PLT) populations. Across all inflammatory conditions studied, recovering patients followed a consistent WBC-PLT trajectory shape that is well-approximated by exponential WBC decay and delayed linear PLT growth. This recovery trajectory shape may represent a fundamental archetype of human physiologic response at the cellular population scale, and provides a generic approach for identifying high-risk patients: 32x relative risk of adverse outcomes for cardiac surgery patients, 9x relative risk of death for COVID-19, and 5x relative risk of death for myocardial infarction.

## Introduction

Trauma, infection, ischemia, and other pathologic conditions trigger an inflammatory response, a set of orchestrated processes involving molecular inducers of cellular and tissue-level effector programs that seek to prevent further injury and to repair existing damage^1,2^. The acute phase of inflammation is typically induced by exogenous molecular inducers from pathogens or endogenous molecules released or exposed following tissue stress or damage^2^. The response is then often mediated by WBCs resident in damaged tissue or by platelets aggregating and activating at the site of injury^1,3^. Homeostatic setpoints are altered temporarily by an acute inflammatory response and return to baseline during resolution^2,4,6^. The identities of many molecular inducers and cellular sensing mechanisms have been established, but response dynamics at the level of WBC and PLT effector populations are poorly understood. For instance, while elevated WBC count is a cardinal sign of acute inflammation, the magnitude of WBC elevation and rates of elevation and resolution associated with favorable acute inflammatory responses are not well-defined^2,4^. Characterization of patient inflammatory state is fundamental to the accurate diagnosis and management of most acute illnesses, but our fragmented understanding of inflammatory responses at the cellular population level limits clinical practice to binarized readouts of serum acute phase proteins and heuristic assessment of blood counts.

We analyzed multivariate temporal relationships in clinical measurements from patients responding to multiple types of trauma associated with surgery, ischemia, and infection. We characterized favorable recoveries and found common dynamics that appear to represent a fundamental unrecognized physiologic program at the cellular population level: uncomplicated recoveries from inflammatory states were defined by exponential decay of WBC and linear growth of PLT. This conserved response pattern may facilitate clinical management of acutely ill patients by providing a personalized benchmark for inflammatory recovery, analogous to the growth curves used to monitor healthy pediatric development^7^.

## Results

### Multivariate inflammatory trajectories

We first considered patients undergoing non-emergency cardiac surgery because this scenario provides (1) a significant inflammatory perturbation with well-defined timing and consistent magnitude (typically involving sternotomy and cardiopulmonary bypass) and (2) relatively stable initial conditions across the patient cohort which excluded those whose procedures were deemed emergency or salvage^8,9^. We studied 4693 patients at Massachusetts General Hospital (MGH): an exploratory cohort with surgery between 01-01-2016 and 30-09-2018, N = 3168, and a validation cohort with surgery between 01-10-2018 and 31-12-2019, N = 1525 (Table 1). Adverse outcomes were defined as death or a post-operative hospital length of stay (LOS) greater than 14 days. We analyzed high-dimensional response trajectories using up to 20 standard protocol clinical laboratory measurements including the complete blood count (CBC) and basic metabolic panel (BMP), before and after surgery (**Fig. 1A**). Unsupervised clustering of these high-dimensional trajectories identified high-risk and low-risk subsets whose mortality rates differed 22-fold (0.8% and 17.8%, supplemental materials 1.2). Multiple individual markers, including WBC and PLT, were highly auto-correlated and cross-correlated (supplemental materials 1.3), suggesting robust co-regulation of blood cell populations and serum markers during the inflammatory response to surgery, and implying that reduced-dimension phenotype definitions would retain sufficient information to identify common features of favorable responses.

**Table 1.**
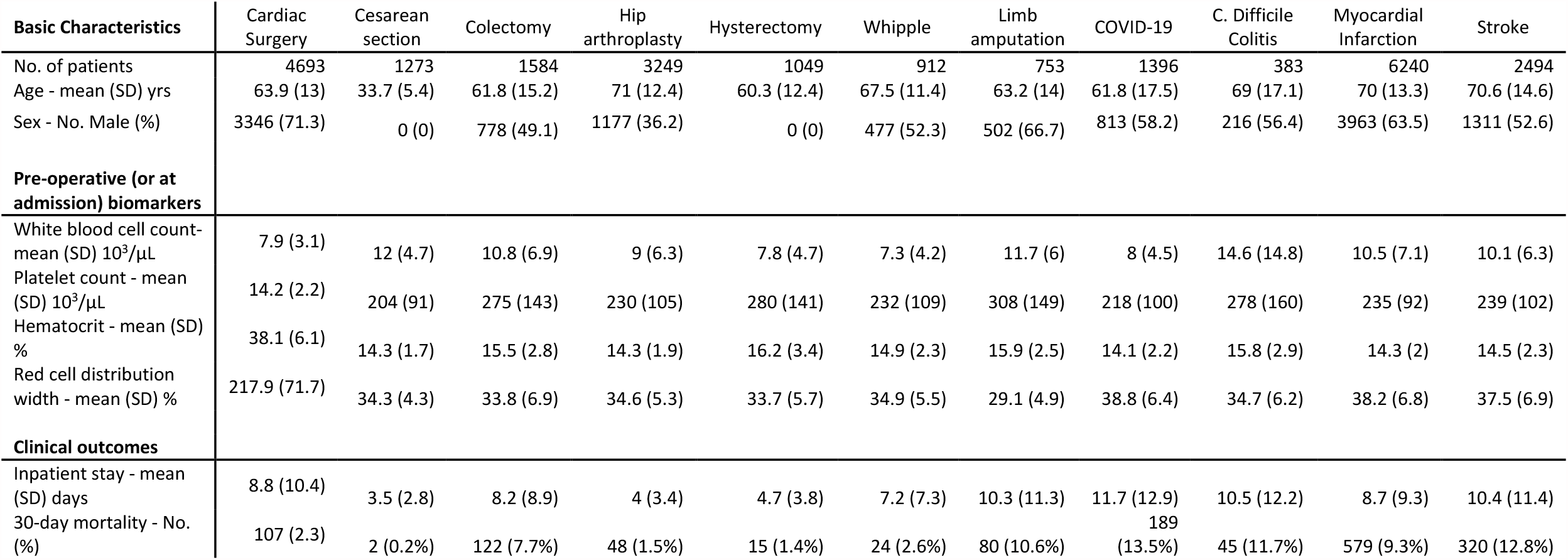
Characteristics of the inflammation cohorts.

**Figure 1.**
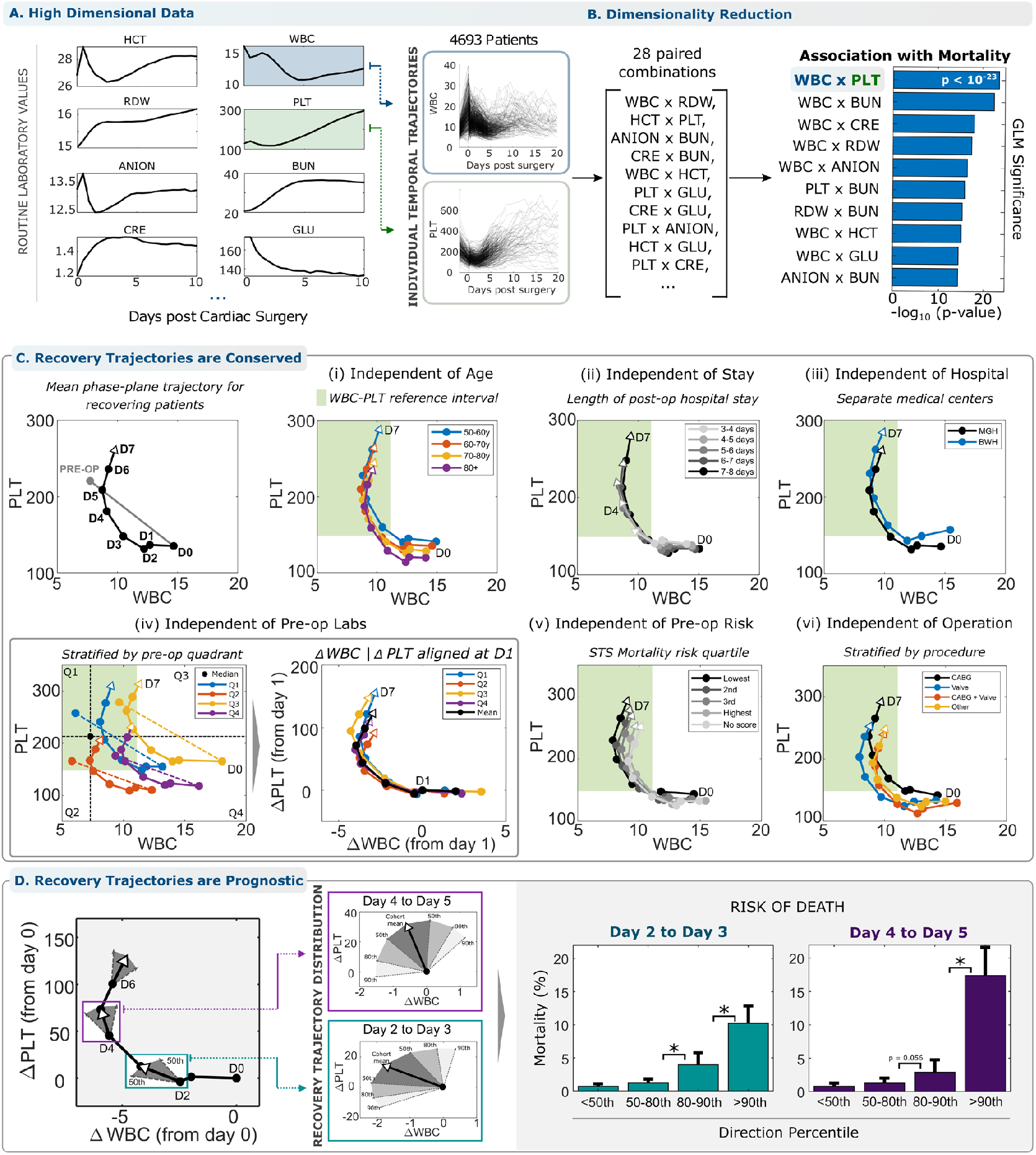
Reduced-dimension analysis of acute inflammatory response in the WBC-PLT phase plane reveals a consistent shape of recovery. **(A)** High dimensional trajectories of cellular and serum markers following cardiac surgery are associated with patient outcome. (**B**) Reducing dimensionality of trajectories retains significant association with patient outcome. The WBC-PLT phase plane retains high association consistent with understanding of inflammatory response mechanisms. (**C**) The average patient with a good recovery (surviving with LOS < 14 days) follows a WBC-PLT trajectory that departs from both WBC and PLT reference intervals (green region) and returns toward the pre-operative baseline over about 5 days. This shape of a good inflammatory recovery persists independent of (i) patient age, (ii) length of hospital stay, (iii) hospital of service, (iv) baseline patient WBC-PLT, (v) pre-op risk defined by the Society of Thoracic Surgeons risk score^30,31^, (vi) cardiac surgery sub-type, patient gender, year of surgery, and general pre-op risk. (See supplemental materials 1.5). (**D)** Deviation from the mean recovery trajectory is prognostic for adverse outcomes. The mean trajectory along with the 50^th^, 80^th^ and 90^th^ percentiles for daily directional changes are shown. Deviation from the direction of the mean trajectory is associated with significant (*, p<0.05) increased risk of death, 14x (CI 8.0-24.1, 0.7% to 10.2%) on day 3 after surgery and 22x (CI: 11.7-39.8, 0.8% to 17.4%) on day 5, between patients below the 50^th^ and above the 90^th^ percentiles. Percentile thresholds were calculated in the exploratory cohort, and outcome rates from the validation cohort. The identification of WBC x PLT as the optimal 2D pair was performed solely using the exploratory cohort. Circles in panels (**D)** denote 1-day time intervals, or from pre-op to immediately post-op.

### WBC-PLT trajectory shape in trauma

Many approaches to dimensionality reduction, such as principal component analysis or t-SNE, may maximize information retention at the expense of interpretability and are optimized for static datasets, rather than time series.^10^ To retain interpretability^11–15^, we performed phase plane analysis, a powerful approach commonly used in engineering and applied mathematics to understand the behavior of dynamical systems by analyzing subsets of system variables as a function of time^16^. We focus on the WBC-PLT plane because WBCs and PLTs play primary roles in inflammation^1,2,6^, and the WBC-PLT combination provided at least as much information about outcomes as other pairwise combinations (**Fig. 1B**, supplemental materials 1.4). Analysis in higher-dimensional phase spaces yielded only incrementally increased associations with outcomes. Patient WBC-PLT trajectories confirm that cardiac surgery is a dramatic inflammatory perturbation, moving more than 95% of patients outside the reference intervals for WBC or PLT (**Fig. 1C**). To investigate whether recovering patients follow homogeneous or heterogeneous paths, we initially looked at the patients with good recoveries: survivors with LOS < 14 days^8,9^. These recovering patients tended to have WBC-PLT trajectories with a consistent shape (**Fig. 1C**). Individual patient trajectories reflect divergent baseline characteristics and variation in clinical events, especially over short periods of time (**Fig. 1A**), but the shape of the average WBC-PLT trajectory nevertheless was conserved when recovering patients were stratified by age, LOS, hospital of service, baseline WBC and PLT, standardized pre-operative risk assessment^8,9^, operation type, patient gender, year of surgery, and more (supplemental materials 1.5). WBC and PLT populations were on average co-regulated in a consistent way during the resolution phase of an effective inflammatory response to cardiac surgery. This unified WBC-PLT recovery shape may provide a benchmark against which individual patient WBC-PLT trajectories can be compared. We thus investigated whether patients whose trajectories closely mimic this shape’s direction have better outcomes. In an independent patient cohort, patients whose trajectory direction diverged from this benchmark had a 14x increased risk of death (CI 8.0-24.1, 0.7% to 10.2%) on day 3 after surgery and a 22x increased risk (CI: 11.7-39.8, 0.8% to 17.4%) on day 5 (**Fig 1D** and supplemental materials 1.6-1.7).

The persistence of this conserved recovery trajectory throughout the cardiac surgical cohort motivated an exploration of these cellular dynamics in patients recovering from trauma associated with a broad range of surgeries for other indications. We analyzed WBC-PLT trajectories for six other surgeries: Cesarean section, colectomy, hip replacement, hysterectomy, limb amputation, and Whipple procedure. See Table 1 and supplemental materials 1.1. **Fig. 2A** shows a similar average recovery shape for all surgeries after adjusting for baseline WBC and PLT and the magnitude of the WBC response. A consistent shape is seen despite the fact that these patient populations are highly diverse in terms of baseline state of health, demographics, and acute indication for surgery. Like the cardiac cohort results in **Fig. 1C**, mean trajectories for these cohorts are consistent when stratified by LOS (supplemental materials 1.8)

**Figure 2.**
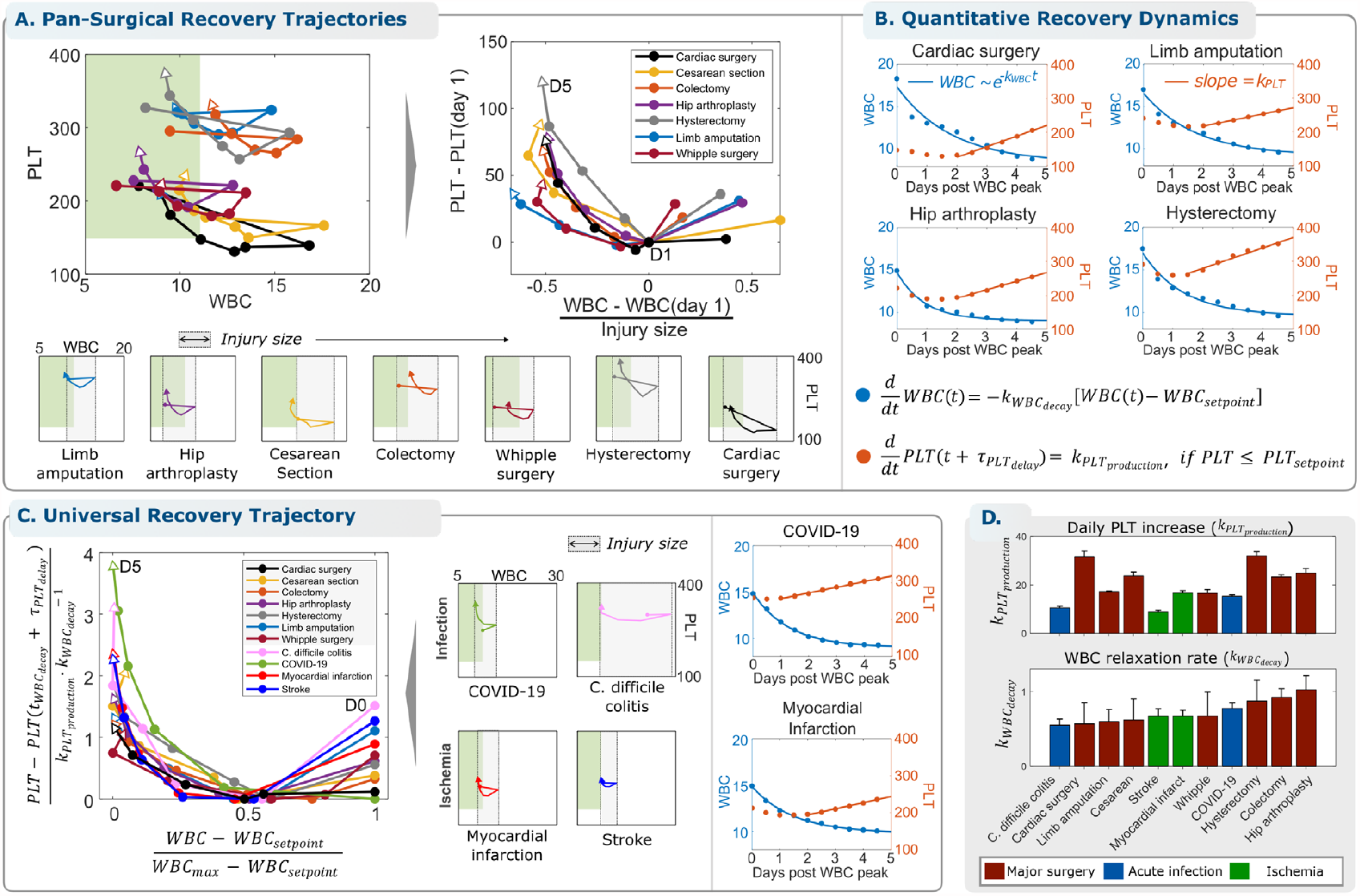
The WBC-PLT recovery shape is conserved across multiple types of traumatic, infectious, and ischemic inflammatory stimuli and demonstrates exponential WBC decay and linear PLT growth. **(A)** Other surgeries are associated with a similar average shape of WBC-PLT recovery. **(B)** WBC dynamics are modeled as simple exponential decay from the maximum WBC, and PLT dynamics are modeled as linear growth starting 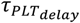 days after the maximum WBC. **(C)** Infectious and ischemic inflammatory insults demonstrate the same average WBC-PLT recovery shape, and a non-dimensionalized^18^ model shows that physiologic response to all inflammatory stimuli considered share the same characteristic features. (D) 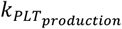 varies from 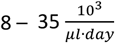 and 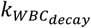 from 0.5 – 1.05 *days*^−1^.

### Exponential WBC decay and linear PLT growth

We found that the WBC dynamics of the typical recovery trajectory could be approximated as an exponential decay starting from the maximum WBC (*WBC*_*max*_) after surgery and falling toward the patient’s pre-inflammation baseline or setpoint (*WBC*_*setpoint*_) with a decay constant of 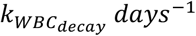 (**Fig. 2B**):

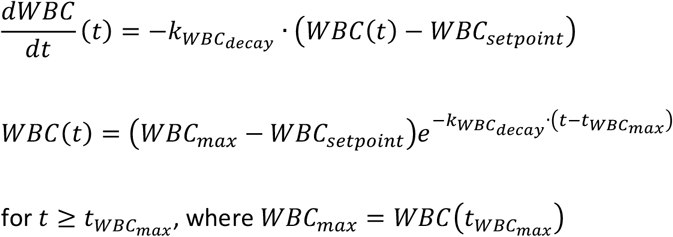

Starting about two days (*τ*_*PLT*_*delay*_) after the maximum WBC 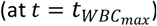, the PLT count of the typical recovering patient grew approximately linearly in time, with a linear growth rate or slope of 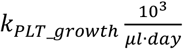 (**Fig. 2B**):

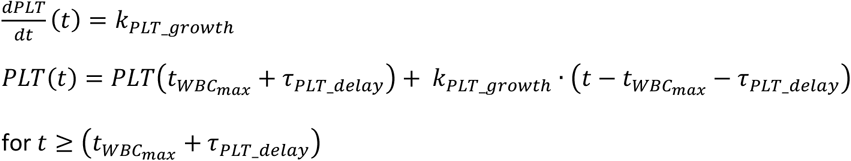

WBC and PLT counts reflect net effects of production, consumption, migration, and turnover, and multiple models of these component processes are consistent with the observed WBC-PLT dynamics. One simple model consistent with this data that is motivated by standard concepts of homeostatic negative feedback control systems^6,17^ couples (i) removal of excess circulating WBCs (i.e., above *WBC*_*setpoint*_) at a constant rate 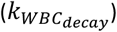 per WBC with negligible introduction of new WBCs into the vascular space and (ii) constant PLT production (*k*_*PLT*_*production*_) with negligible consumption starting about 2 days later. Both processes continue at least until reaching the patient’s baseline setpoints. According to this model, excess WBCs are removed at random after circulating for an average duration of ∼1-2 days *k*_*WBC*_*decay*_^−1^, with a standard deviation of 1-2 days (*k*_*WBC*_*decay*_^−1^), corresponding to a half-life of 0.7 – 1.4 days. The PLT growth is synchronized to start about two days (*τ*_*PLT*_*delay*_) after the time of maximum WBC and continues until the PLT count has reached or exceeded the patient’s baseline *PLT*_*setpoint*_. In the cardiac surgery cohort where more data was available, most recovering patients appear to overshoot their baseline *PLT*_*setpoint*_(**Fig. 1C**), consistent with the hypothesis of feedback delay (*τ*_*PLT*_*delay*_) and other mechanisms. See supplemental materials 1.9 for more detail.

### Recovery in infection and ischemia

Given the resilience of the conserved recovery trajectory shape across a diverse range of surgical trauma, we investigated whether the pattern persists in non-traumatic inflammatory conditions: two types of infection (COVID-19 and Clostridium difficile colitis) and two types of ischemia (myocardial infarction and stroke). See Table 1 and Methods for cohort details. **Fig. 2C** shows that the same basic response shape is identified and that WBC and PLT kinetics for the average recovering patient in each cohort are well-explained by exponential WBC decay and linear PLT growth. As in the surgical cohorts, this characteristic response is maintained when the cohorts are stratified by gender, age, LOS, and baseline WBC-PLT (supplemental material 1.10). To reveal characteristic features of WBC and PLT dynamics intrinsic to all inflammatory stimuli, we non-dimensionalized^18^ the model using the following dimensionless variables (*w, p, t*^*\**^):

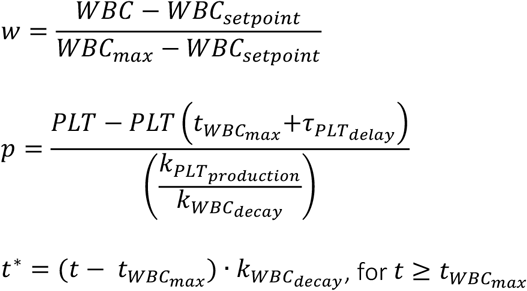

**Fig. 2C** shows that good inflammatory recoveries from all traumatic, infectious, and ischemic stimuli considered can be well-approximated by the following simple mathematical model:

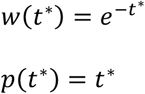

For many inflammatory conditions, 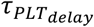is approximately equal to the half-life for WBC decay, corresponding to *(w,p) = (0*.*5,0)* in Fig. 2C.

### Trajectory-based patient risk assessment

**The analysis in Fig. 1D** shows that directional deviation from this conserved WBC-PLT recovery shape is associated with elevated risk of adverse outcomes following cardiac surgery. WBC-PLT position can also be considered to provide an integrated trajectory-based risk model. Illustrating this approach to individual patient risk assessment, the left panel of **Fig. 3A** plots the hospital course of a 55-year-old female with a prior mechanical aortic valve replacement and high STS pre-operative risk of mortality (PROM)^8,9^ (10%, 95^th^ percentile). Repeat sternotomy for aortic valve replacement was complicated by intra-operative bleeding and hypotension requiring rapid transfusion and resuscitation, but the post-op recovery was otherwise smooth. Daily comparison of the patient’s WBC-PLT position and direction yields a high initial risk that steadily declines as her WBC normalizes, and her trajectory merges quickly with the reference shape of recovery. In contrast, the right panel of **Fig. 3A** illustrates the clinical course for an 84-year-old female with a history of angina and moderate STS pre-operative mortality risk (2.6%, 75^th^ percentile) who initially recovered well after successful three-vessel coronary artery bypass grafting. Counts normalized by post-op day 4, but the rising WBC and subtly declining PLT thereafter nevertheless correspond to a marked deviation from the favorable trajectory and a sharp rise in directional risk. On post-op day 6, she developed fever and shortness of breath leading to a diagnosis of pulmonary embolism and initiation of heparin. A precipitous interval decline in PLT on day 7 led to diagnosis of heparin-induced thrombocytopenia, intensive care unit readmission, and a subsequent prolonged hospital stay of one month. These two cases illustrate how the WBC-PLT recovery shape can be used to identify high-risk patients who recover smoothly, as well as to detect smoldering issues which precede adverse events. For the cardiac surgery cohort in general, patients outside the 80^th^ percentiles of position and direction have a relative risk of adverse outcomes of 32 on day 3 (CI: 13-79, 1.4% to 44%) and 33 on day 4 (CI: 15-75, 1.6% to 53%). Extending this analysis to the other inflammatory scenarios studied revealed significant relative risks: 9.2 (2.7-31.2, 4.3% to 40%) for death from COVID-19, 4.9 (3.1-7.6, 5.8% to 28%) for death following myocardial infarction, and 10.1 (CI: 4-26, 5% to 50%) for hip replacement, with all results from out-of-sample testing (Fig. 3C). In the validation cardiac surgery cohort, 97% (29/30) of deaths occurred in patients whose day 4 position or direction were above the 80^th^ percentile, with 83% (25/30) occurring in patients with both position and direction above the 80^th^ percentile. Corresponding specificities were 71% (865/1226) and 92% (1125/1226). See supplemental materials 1.13 for more detail.

**Figure 3.**
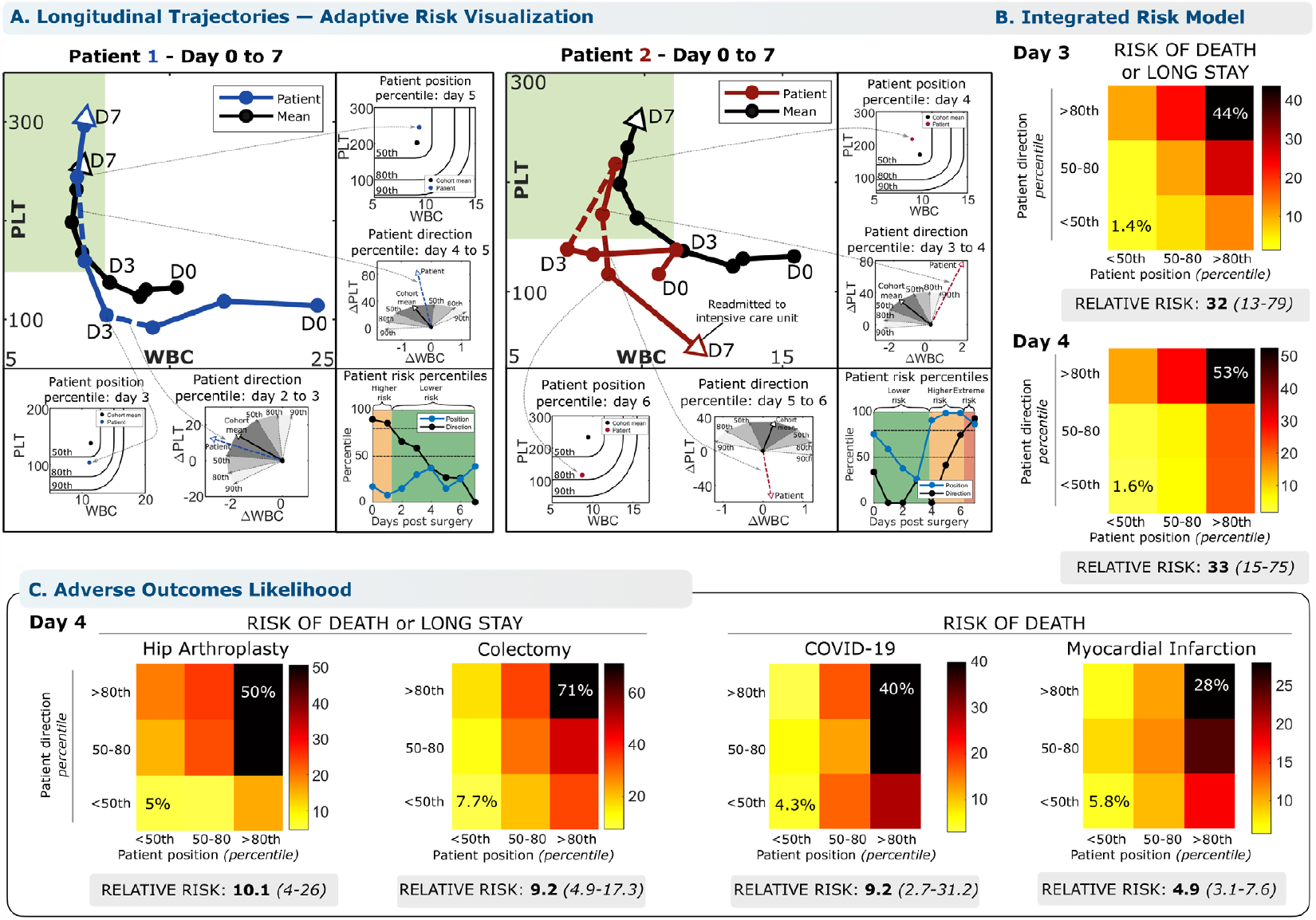
The conserved WBC-PLT trajectory shape provides a generic benchmark for personalized patient risk assessment. WBC-PLT position and direction provide an integrated trajectory-based risk model. Two patient examples are provided, one involving recovery of an initially high-risk patient (**A**, Patient 1), and the other involving deterioration of an initially stable patient (**A**, Patient 2). See main text for case details. (**B**) Across the entire validation cohort, the trajectory-based assessment identified patients at high risk of mortality and prolonged length of stay (LOS). Position and direction jointly contribute to risk, with patients outside the 80^th^ percentile for both position and direction having elevated likelihood of poor outcomes. Compared to patients with good position and direction (<50^th^ percentile), patients with poor position and direction (>80^th^ percentile) on post-op day 4 had a 33-fold increased relative risk of adverse outcomes (CI: 13-79, 53% vs 1.6%) and stayed an average of 13 days longer in hospital. (**C**) For other inflammatory stimuli studied, positional and direction deviation were similarly associated with significant increases in relative risks of adverse outcomes for patients with sufficient data available: 10x for adverse outcomes following hip replacement, 9x for colectomy, 9x for mortality from COVID-19, and 5x for mortality from myocardial infarction. See Methods for more detail. Reference percentiles for position and direction and outcome likelihoods across the first 7 days post-op are given in the supplemental materials 1.13. Position and direction thresholds were calculated from the exploratory cohort, and outcome rates from the validation cohort. Position percentiles were calculated treating WBC below and PLT above the mean as non-contributing.

## Discussion

We identified a conserved shape of human acute inflammatory recovery defined by co-regulation of WBC and PLT populations. The shape spans diverse traumatic, infectious, and ischemic inflammatory stimuli, irrespective of pre-inflammatory patient state and risk, and is robust across multiple years of clinical care and two different hospitals of service. This WBC-PLT trajectory shape thus appears to represent a fundamental unrecognized dynamic mechanism of human physiology.

The WBC-PLT trajectory shape identifies patients with 5-33x relative risk of adverse outcomes in our study cohorts and may provide a generic personalized benchmark for tracking individual patient recovery, analogous to a pediatric growth-chart^19^. Complications due to additional inflammatory stimuli or other pathologic events are expected to perturb the benchmark shape or superimpose other dynamics. Recognition of this WBC-PLT shape as the hallmark of a favorable response will also help guide investigation of the molecular and cellular signals driving functional and dysfunctional inflammatory responses. The robustness of this recovery shape shows that healthy inflammatory responses are similar, in contrast to those associated with adverse outcomes (supplemental materials 1.6) which can be highly divergent. In general, this contrasting homogeneity and heterogeneity for successful and unsuccessful outcomes has been identified in other biological systems as the Anna Karenina Principle.^20^

The mathematical characterization of inflammatory recovery as exponential WBC decay toward baseline with delayed linear PLT growth toward baseline is consistent with conceptual models of homeostatic control of WBC and PLT relative to their setpoints^2,6,17^. Modeled *k*_*PLT*_*production*_ varies significantly with inflammatory stimulus, but 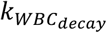 does not, despite large variation in WBC. The range of 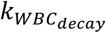(0.5 -1.0 days^-1^) is similar to experimentally observed neutrophil turnover rates and monocyte clearance times in myocardial infarction^21^ and other settings^22^. The PLT growth ∼1-2 days 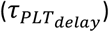 after peak WBC suggests that hemostasis is achieved shortly after pro-inflammatory signals have peaked. Recoveries for many inflammatory conditions appear to overshoot, reaching *PLT*(*t*) > *PLT*_*setpoint*_ (**Fig. 2B and 2C**). Future investigations are needed to validate this observation and more generally to define the logic of the hypothesized control systems, dissecting the contributions of production, consumption, senescence, and other component processes. Future work is also needed to understand how each patient’s pre-operative WBC-PLT setpoint is stored to enable consistent recovery toward that baseline.

All major results were validated in out-of-sample testing or using cross-validation approaches. Results for the cardiac surgery cohort were replicated in separate cohorts both from the same hospital during different time periods and from a different hospital in the same network, and in subset analyses controlling for surgery type, and pre-operative biomarkers and risk profiles. However, this study was limited to retrospective analysis, and some patient cohorts were significantly restricted by required minimum data availability. Follow-up prospective data with higher-frequency measurements are needed to define these responses more precisely and to determine whether interventions guided by this conserved recovery shape may lead to improved patient outcomes. Future work is also warranted to determine whether the shape of the effective inflammatory response we have identified is present in other inflammatory settings such as malignancy, autoimmunity, and the resolution of chronic inflammation.

## Methods

### Clinical data collection

Data was initially collected from all adult Massachusetts General Hospital (MGH) patients who underwent cardiac surgery between 01-01-2016 and 31-12-2019 (N=6054). Data was gathered using the Partners Healthcare Research Patient Data Registry (RPDR) and Electronic Data Warehouse (EDW) under a research protocol approved by the Partners Healthcare Institutional Review Board. For each patient, all blood count (CBC) and basic metabolic panel (BMP) measurements during the surgery-associated hospital stay were collected. Patient demographics, risk factors, surgical factors and post-operative outcomes were collected from a dataset adjudicated by the Massachusetts General Hospital Division of Cardiac Surgery for contribution to the national Society for Thoracic Surgery (STS) database. Patients were excluded if their surgery was designated as a salvage or emergency procedure, if they had a post-operative stay of less than 48hrs, or if they did not have at least 1 valid CBC and BMP post-surgery, giving a cohort of N = 4693. Cardiac surgeries were divided into four groups: isolated coronary artery bypass graft (CABG), valve surgeries (aortic valve replacement, or mitral valve replacement/repair), CABG + valve, or other. An independent cohort was derived from cardiac surgeries at Brigham and Women’s Hospital (BWH) between Jan-01-2016 and Sep-30-2019 (N = 1988). To minimize potential bias^23^, for initial analysis only data from MGH surgeries between 01-01-2016 and 30-09-2018 (N=3168) were compiled (denoted the exploratory cohort). All exploratory data analysis was performed on this cohort. Following finalisation of the analysis plan, data from surgeries between 01-10-2018 and 31-21-2019 (N=1525) was compiled (denoted as the validation cohort) and analysed.

Following analysis of the cardiac surgery cohort, clinical data was collected from all adult MGH patients who underwent any of 6 other major surgeries (colectomy, Cesarean section, hip arthroplasty, hysterectomy, limb amputation, Whipple surgery), or who were diagnosed with any of 4 acute inflammatory conditions (myocardial infarction - MI, stroke, infectious colitis, or novel coronavirus infection [COVID-19]), between 01-01-2016 and 31-12-2020. Patients with associated hospital stays of less than 2 days and those undergoing laparoscopic procedures were excluded. If a patient had multiple visits associated with the same surgery type or inflammatory diagnosis between 2016 and 2020, only the earliest visit was included. As with the cardiac surgery cohort, all CBC and BMP measurements associated with the hospital stay were collected, along with demographics, admit and discharge dates, and all-cause mortality. Cohort sizes pre- and post-exclusion, and any exclusion criteria specific to each sub-cohort are presented in supplemental materials 1.1. Following exclusions, each cohort was split evenly, with the earliest half (by diagnosis or surgery date) taken as the exploratory set, and the latter half taken as the validation set. Unless otherwise noted, all prediction-based results in the manuscript were calculated out-of-sample, with thresholds defined using exploratory sets, and performance measured in the validation sets.

From the CBC and BMP, 20 biomarkers were considered (reference intervals at MGH given in supplemental materials 1.14). Results for the estimated glomerular filtration rate (eGFR) were often binarized when above 60mL/min/1.73m^2^, and as such these results may not be fully representative. Except for the white cell count (WBC), none of these biomarkers is typically considered a direct measure of inflammatory state. However, all are routinely used in assessment of both acute and chronic inflammation^24–28^. Primary outcomes for this study were 30-day (post-discharge) all-cause mortality and length of hospital stay (LOS). LOS was calculated as the time between initial incision and discharge for surgical cohorts, and between initial hospital admission and discharge for non-surgical cohorts. For surgeries with mean LOS > 7 (amputation, cardiac, colectomy, Whipple), a “long stay” was defined as LOS > 14, while for surgeries with LOS < 7, long stay was defined as LOS > 10.

### Identification of optimal biomarker pairs

For initial analysis, clinical laboratory test results were interpolated and evenly sampled every 12 hrs post-operation. Patient response clusters in the cardiac exploratory cohort were derived using k-means clustering^29^ applied to patient CBC and BMP measurements throughout the surgery-associated hospital stay (full details and results in supplemental materials 1.2). This process highlighted 8 of the 20 biomarkers which most strongly stratified patient responses: anion gap (ANION), blood-urea nitrogen (BUN), creatinine (CRE), hematocrit (HCT), glucose (GLU), platelet count (PLT), red cell distribution width (RDW), and WBC. Dimensionality was further reduced by identifying the optimal pair of biomarkers (from the 28 possible combinations) for risk stratification. High-performance pairs were identified by considering the significance (p-value, *χ*^2^ test) of a generalized linear model predicting mortality using the interpolated post-op day 1 measurements from each biomarker pair. Clinical and physiologic interpretability were also subjectively considered, supporting the choice of WBC-PLT as the chosen 2D distillation of the high-dimensional dataset.

### White blood cell (WBC) – platelet (PLT) phase-plane analysis in cardiac surgery

WBC-PLT dynamics post cardiac surgery were analysed by considering joint movement of a patient’s WBC and PLT counts in a 2D phase-plane. Mean WBC-PLT trajectories were calculated over the first week post-op, using each patient’s interpolated WBC and PLT trajectories. Mean responses were calculated for a wide variety of sub-cohorts: stratified by outcome, LOS, age, cardiac surgery type, pre-operative WBC and PLT values, pre-operative mortality risk (calculated through the STS risk score calculator^30,31^), and in both the MGH and BWH cohorts. The favourable inflammatory response was defined as the mean WBC-PLT trajectory for all cardiac surgery patients who survived and did not have a long stay (LOS > 14).

Given WBC-PLT data is not available after a patient is discharged, the number of patients used to define the favourable response trajectory decreases over time and maybe create a bias if patients who are discharged on a given day have WBC-PLT values significantly above or below the cohort mean. However, as illustrated in **Fig 1C**, this bias does not appear to alter the shape of the recovery curve, with the shape being invariant when limited to patients within more narrow LOS distributions.

### Analysis of WBC and PLT dynamics in broader inflammatory settings

From analysis of the cardiac surgery cohort, it was hypothesized that following an isolated acute inflammatory injury, WBC would decay exponentially, while PLT would rise linearly. To test this hypothesis, linear and exponential models were fit to the mean WBC and PLT trajectories for the other 10 inflammation cohorts. To account for differences in precise timing of the acute inflammatory response, patients were aligned based on the timing of their maximum WBC count in the 3 days post-operation (surgical cohorts), 3 days post admission (ischemia cohorts), or over their entire stay (infection cohorts). The alternative alignment for the infection cohort was chosen to reflect the fact that viral infections represent a slower-onset acute event than surgical trauma or ischemia, and thus the acute inflammatory response may not necessarily occur near the time to diagnosis. An exponential decay model (*y* = *a* + *be*^−*ct*^; parameters: a, b, c) and a linear model (*y* = *d* + *e * t*; parameters: d, e) were fit to the mean WBC and PLT counts over the first 5 days post-alignment. The PLT model was fit with a lag parameter *τ*, which accounts for potential delay between peak WBC and the start of PLT growth. Models were fit using *MATLAB’s* statistical toolbox, to the mean post-peak WBC and PLT curves interpolated and evenly sampled every 12hrs. For parameter estimation, patients who did not have at least 5 days of post-peak measurements available were excluded.

For each cohort, the response trajectory (**Fig 2**) was calculated as the mean WBC-PLT response for all patients who did not experience adverse outcomes (surgical: mortality or long stay; non-surgical: mortality), and who had at least 5 days of WBC and PLT measurements post-operation (surgical cohorts), post admission (ischemia cohorts), or post peak WBC count (infection cohorts). Patients who did not have a WBC increase of at least 2 units from pre-op to post-op (surgical cohorts), from admission to peak (ischemia and infection cohorts) were excluded from this calculation due to expected challenges distinguishing analytic from biologic variation. The ischemia and infection cohorts had their WBC-PLT trajectories aligned using the same process as outlined above for the exponential and linear model fitting. To allow for comparison of responses, PLT was normalized by the model-estimate of daily PLT increase (k_PLT_PRODUCTION_), and WBC was normalized by injury size. For surgical cohorts, the injury size was estimated as the difference between mean pre-op and immediate post-op WBC count. For non-surgical cohorts the admission WBC may not meaningfully reflect baseline WBC, so instead, injury size was calculated as the difference between peak WBC count and baseline WBC as estimated from the exponential decay model.

### WBC-PLT risk analysis

For all surgeries, patient risk of adverse outcomes (death or long stay) on a given day was calculated by comparing patient position and movement direction in the WBC-PLT phase-plane to the benchmark inflammatory response trajectory for that cohort (defined as the mean WBC-PLT response of patients in the exploratory cohort without adverse outcomes). Positional risk was calculated as the distance from the mean trajectory, after normalizing WBC and PLT by pre-operative means, and treating WBC below, and PLT above the mean as not contributing to distance.

Directional risk was calculated as the angle of a patient’s normalized daily WBC and PLT change vector against the normalized mean trajectory WBC-PLT change vector. For ease of interpretation, positional and directional risk were converted to percentiles, based on data from the exploratory cohorts. For risk calculations all percentiles and thresholds were calculated in the exploratory cohort, while all risk stratifications using these thresholds were calculated in the validation cohort. Choices of percentile thresholds in **Fig 3** were made from analysis of the cardiac surgery exploratory cohort only.

For the infectious disease cohorts (COVID-19, C. diff. colitis), there may be a delay between the timing of diagnosis, and the onset of the acute injury. Risk calculations in these cohorts were performed after alignment of the patient cohort based on timing of the maximum WBC value within the first 72hrs post admission. Risk stratification remained significant independent of this alignment (supplemental materials 1.12)

Equivalent analysis of other (high-significance) biomarker pairs beyond WBC-PLT was also performed (supplemental materials 1.4). Risk calculations were performed using interpolated and evenly sampled WBC and PLT values, which may introduce mild data leakage – where interpolation introduces future information before it would be available clinically. We investigated this possibility by recreating key results using non-interpolated lab values and found similar degrees of risk stratification (supplemental materials 1.15).

### Statistical analysis

All statistical analysis was performed in *MATLAB*. For all continuous variables, unless otherwise noted, we report means and use analysis of variance (ANOVA) for population comparisons. For categorical variables we report percentages and use a chi-square test for population comparisons. Statistical significance of trajectory clusters was tested through comparison to synthetic data, using a method outlined by Liu et al^29^. Thresholds for statistical significance were set at p = 0.05. For event rates, confidence intervals were calculated assuming binomial distributions. Statistical methods for results presented in the supplemental materials are described within those materials.

## Supporting information

STROBE Checklist

## Data Availability

Summary data that is not presented in tables, figures or supplemental materials is available from the authors upon reasonable request. Due to IRB restrictions on sharing of protected health information, individual patient data cannot be made available.

## Acknowledgements

We thank Rahul Deo and Erik Reinertsen for helpful feedback, Chin Siang Ong and Steven Song for help with access and orientation to datasets, David Louis for helpful discussions, and the Partners Healthcare Research Patient Data Registry group for facilitating use of their database. Portions of the computational analysis in this study were conducted on the O2 High Performance Compute Cluster at Harvard Medical School.

## Ethics Statement

The study protocol was approved by the local institutional review board (IRB) at Massachusetts General Hospital.

## Author contributions

ADA, BHF, JCTC, JMH designed the study, analyzed data, and wrote the manuscript. ADA, BHF, JCTC, JMH, and TS contributed to data collection, data interpretation, and manuscript editing.

