## Supplemental Materials for "White Blood Cell and Platelet Dynamics Define Human Inflammatory Recovery"

**SUPPLEMENTAL MATERIALS 1**

1.1 **Cohort exclusion protocols**

Following analysis of the cardiac surgery cohort, clinical data was collected for 10 other acute inflammation cohorts, 6 major surgeries (Cesarean section, colectomy, hip arthroplasty, hysterectomy, limb amputation, Whipple surgery), 2 ischemic cohorts (myocardial infarction and stroke), and 2 infection cohorts (COVID-19, and Clostridium difficile colitis). Patients were excluded if they were under 18yrs old, had a less than 2 day associated hospital stay, or underwent a laparoscopic procedure (for surgical cohort). Some cohorts had additional inclusion/exclusion criteria specific to that setting:

- Amputation: Surgeries were only included if they involved amputation of a leg (above or below knee), arm (above or below elbow), or of a whole foot or hand. Amputations of fingers or toes were not included.
- Colectomy: Colectomy was defined as any major (invasive, non-laparoscopic) surgery of the small or large bowel, predominantly small bowel resection or full or partial large bowel resection/colectomy.
- Stroke: Stroke was defined as any diagnosis of a stroke or cerebrovascular accident.
- C. difficile colitis: The colitis cohort was limited to patients with a diagnosis of C. difficile colitis or infectious colitis, or with a diagnosis of colitis, and a confirmed positive c difficile toxin assay.

The cardiac surgery cohort was collated from a manually curated dataset adjudicated by the Massachusetts General Hospital Division of Cardiac Surgery for contribution to the national Society for Thoracic Surgery (STS) database. For all other cohorts, data was collected by filtering electronic health record databases for keywords associated with the surgery or diagnosis. For each cohort, terms were selected based on author clinical experience. For each cohort, a random sample of patient health records were manually checked to ensure that the database listed diagnosis/procedures accurately reflected information in their medical charts. Due to the nature of the Partners Healthcare network databases, a small number of patients in each (non-cardiac surgery) cohort may not have fully received treatment at MGH, instead receiving part of their treatment at one of the other hospitals in the Partners Healthcare network.

In addition to total cohort exclusions above, for accurate calculation of trajectories and model parameter estimation in **Fig 2**, patients with 5 days of post-WBC peak data or more were included. To focus on situations where biological variation could be confidently distinguished from analytic variation, patients who did not have an inflammation-induced WBC increase of at least 2 units were excluded.

The size of each of the cohorts after each exclusion are included in **eTable 1.**

**eTable 1 – Cohort sizes after various exclusions**

|  |  | Cohort for overall analysis |  |  | Cohort for Fig 2 trajectories |
| --- | --- | --- | --- | --- | --- |
| Cohort | Initial size | No repeat visits and LOS ≥ 2 | $\Delta$WBC ≥ 2 | Post-peak LOS ≥ 5 | Survivor |
| Limb amputation | 1478 | 753 | 229 | 133 | 87 |
| Colectomy | 2571 | 1584 | 472 | 243 | 165 |
| Cardiac surgery | 4693 | 4693 | 2812 | 1836 | 1465 |
| Cesarean section | 15682 | 1273 | 612 | 49 | 39 |
| Hip arthroplasty | 9085 | 3249 | 496 | 142 | 100 |
| Hysterectomy | 2588 | 1049 | 237 | 68 | 51 |
| Whipple surgery | 1053 | 912 | 136 | 70 | 49 |
| COVID-19 | 1686 | 1396 | 628 | 119 | 100 |
| C. Difficile colitis | 634 | 383 | 117 | 54 | 46 |
| Myocardial Infarction | 8132 | 6240 | 1262 | 656 | 327 |
| Stroke | 12889 | 2494 | 513 | 287 | 103 |

**1.2 Clustering of high-dimensional cardiac surgery clinical measurements**
To identify potential patterns in the high-dimensional CBC and BMP data in the exploratory cardiac surgery cohort, unsupervised learning methods were used. Patient response clusters were derived using k-means clustering applied to patient CBC and BMP measurements throughout their surgery-associated hospital stay. Measurements were normalized (by pre-operative means), interpolated and sampled every 12hrs until discharge, with post-discharge values set to 0. The number of clusters (5) was the maximum number which resulted in all groups having more than 50 patients. Patients with fewer than 3 CBC and BMP measurements were not included when defining clusters but were assigned to their nearest group afterwards. Clinical tests which showed insignificant (<10%) variation across clusters were excluded, leaving 10 measurements – anion gap (ANION), blood-urea nitrogen (BUN), creatinine (CRE), hematocrit (HCT), hemoglobin (HGB), glucose (GLU), platelet count (PLT), red cell distribution width (RDW), red cell count (RBC) and WBC. Given the extremely high correlation between HCT, HGB and RBC, only HCT was included in the final clustering, leaving 8 measurements.

The mean trajectories for the five clusters are illustrated in **eFigure 1,** alongside surgery mixes, and cluster associations with post-operative outcomes, with summary characteristics given in **eTable 2**. Despite a ~20-fold stratification of mortality risk from lowest to highest cluster (0.8% to 18%), the clusters show qualitatively similar behaviour, with main differences occurring in pre-operative mean, and the speed at which dynamics occur. This robust pattern suggests the existence of more fundamental unified inflammatory response, and motivates further distillation of the results in lower dimensional spaces.

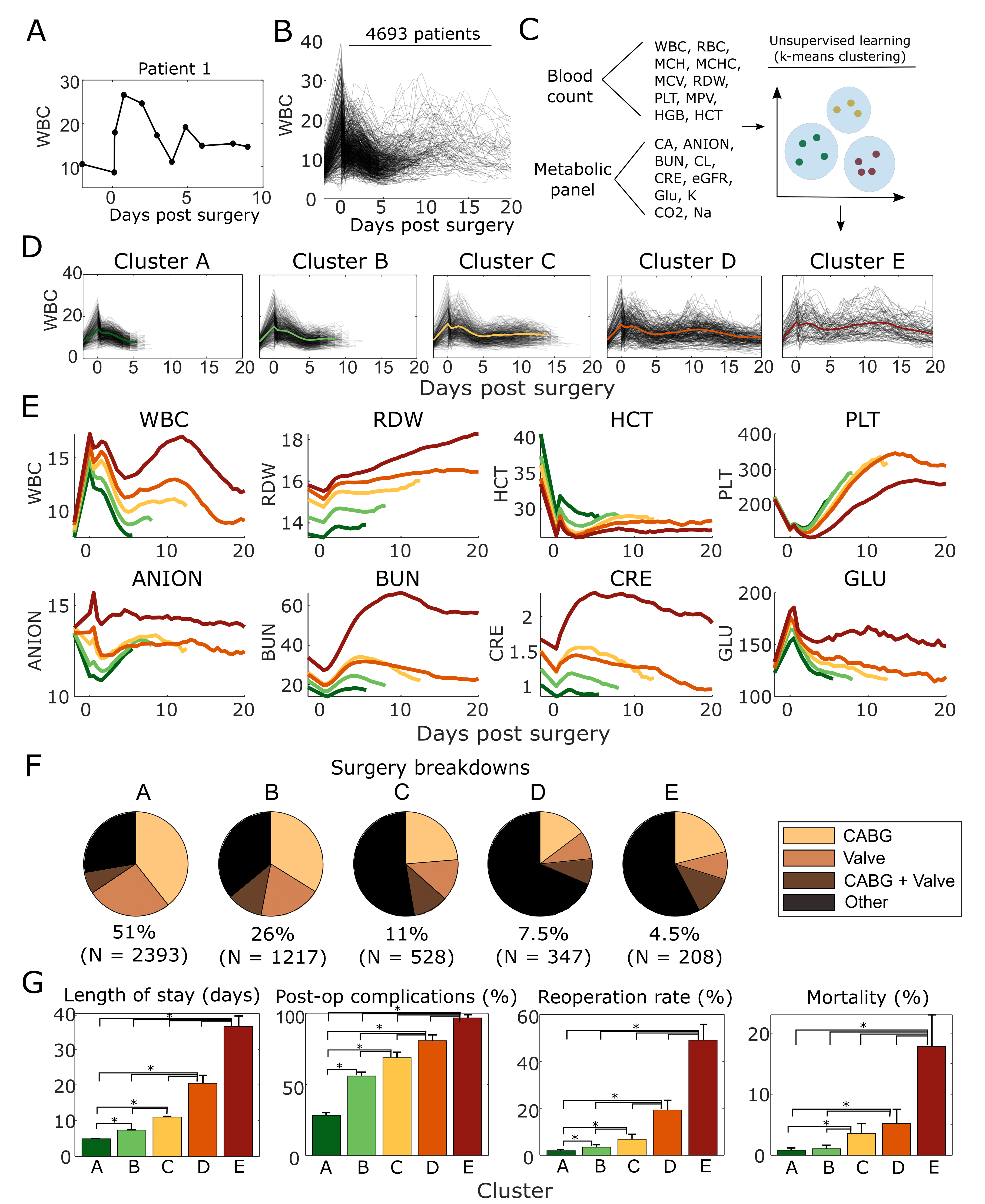

**eFigure 1 – High-dimensional clusters of response to cardiac surgery defined from routine clinical laboratory tests.** Individual lab test results were interpolated (A), and these individual test trajectories (B) were considered for 20 standard complete blood count (CBC), basic metabolic panel (BMP), and other clinical laboratory tests (C). K-means clustering identified five distinct clusters (D, E), associated with significant differences (p<0.05) in surgery type (F) and post-operative outcomes (G). Day 0 corresponds to the first blood count measurement post-surgery, while day -2 corresponds to pre-operative measurements. * denotes values in (G) that are statistically significantly different (p > 0.05) from each other.

**eTable 2 – Cardiac surgery high-dimensional cluster characteristics**

| **Basic Characteristics** | Total | Cluster A | Cluster B | Cluster C | Cluster D | Cluster E |
| --- | --- | --- | --- | --- | --- | --- |
| No. of patients (%) | 4693 (100%) | 2393 (51%) | 1217 (26%) | 528 (11%) | 347 (7.5%) | 208 (4.5%) |
| Age - mean (SD) yrs | 63.9 (13) | 62.3 (12.7) | 66.4 (13) | 65.7 (12.7) | 62.1 (14.4) | 66.4 (12) |
| Sex - No. Male (%) | 3344 (71.3) | 1778 (74.3) | 854 (70.2) | 360 (68.2) | 209 (60.2) | 143 (68.8) |
| Race - No. White/Caucasian (%) | 4047 (86.2) | 2093 (87.5) | 1063 (87.3) | 442 (83.7) | 275 (79.3) | 174 (83.7) |
| Major category surgeries - No. (%) | 2959 (63.1) | 1732 (72.4) | 780 (64.1) | 250 (47.3) | 109 (31.4) | 88 (42.3) |
| **Pre-operative risk factors** |  |  |  |  |  |  |
| Smoking history - No. (%) | 2508 (53.4) | 1175 (49.1) | 715 (58.8) | 306 (58) | 188 (54.2) | 124 (59.6) |
| Diabetic - No. (%) | 1359 (29) | 555 (23.2) | 406 (33.4) | 174 (33) | 120 (34.6) | 104 (50) |
| Lung disease - No. (%) | 680 (14.5) | 209 (8.7) | 196 (16.1) | 109 (20.6) | 98 (28.2) | 68 (32.7) |
| Dyslipidemia - No. (%) | 3589 (76.5) | 1776 (74.2) | 968 (79.5) | 417 (79) | 255 (73.5) | 173 (83.2) |
| Endocarditis - No. (%) | 267 (5.7) | 71 (3) | 82 (6.7) | 47 (8.9) | 40 (11.5) | 27 (13) |
| Family history of coronary artery disease - No. (%) | 562 (12) | 339 (14.2) | 126 (10.4) | 53 (10) | 27 (7.8) | 17 (8.2) |
| Hypertension - No. (%) | 3643 (77.6) | 1779 (74.3) | 992 (81.5) | 432 (81.8) | 258 (74.4) | 182 (87.5) |
| Pulmonary vascular disease - No. (%) | 560 (11.9) | 188 (7.9) | 179 (14.7) | 87 (16.5) | 56 (16.1) | 50 (24) |
| Renal Failure - No. (%) | 89 (1.9) | 3 (0.1) | 22 (1.8) | 28 (5.3) | 19 (5.5) | 17 (8.2) |
| **Pre-operative biomarkers** |  |  |  |  |  |  |
| White blood cell count- mean (SD) 10^3^/µL | 7.9 (3.1) | 7.5 (2.2) | 8 (3.1) | 8.2 (2.9) | 8.7 (5.3) | 9 (3.4) |
| Red cell distribution width - mean (SD) % | 14.2 (2.2) | 13.5 (1.3) | 14.3 (2) | 15.1 (3) | 15.6 (2.5) | 15.8 (3) |
| Hematocrit - mean (SD) % | 38.1 (6.1) | 40.5 (4.8) | 37.4 (5.9) | 36.2 (6.3) | 34.3 (6.7) | 33.5 (6.4) |
| Platelet count - mean (SD) 10^3^/µL | 217.9 (71.7) | 216.7 (61.9) | 221.9 (77.5) | 216.6 (73.6) | 216.5 (85.4) | 214.5 (86.2) |
| Anion gap - mean (SD) mmol/L | 13.5 (2.5) | 13.4 (2.5) | 13.5 (2.4) | 13.8 (2.7) | 13.5 (2.4) | 13.8 (2.6) |
| Blood-urea nitrogen - mg/dL | 22.4 (12.1) | 18.6 (7.2) | 22 (10.7) | 26.2 (14.9) | 25.2 (14.3) | 33.7 (18.2) |
| Creatinine - mean (SD) mg/dL | 1.2 (0.9) | 1 (0.3) | 1.2 (0.8) | 1.5 (1.3) | 1.5 (1.3) | 1.7 (1) |
| Glucose - mean (SD) mg/dL | 126.2 (41.9) | 122.7 (39.3) | 127 (41.9) | 129.6 (44.8) | 127.1 (46.9) | 133.3 (41.6) |
| **Post-operative outcomes** |  |  |  |  |  |  |
| Inpatient stay - mean (SD) days | 8.8 (10.4) | 4.9 (2.3) | 7.4 (1.2) | 11 (2.1) | 20.5 (20.9) | 36.4 (21.4) |
| 30-day mortality - No. (%) | 107 (2.3) | 20 (0.8) | 13 (1.1) | 19 (3.6) | 18 (5.2) | 37 (17.8) |
| Renal failure - No. (%) | 140 (3) | 13 (0.5) | 9 (0.7) | 24 (4.5) | 14 (4) | 80 (38.5) |
| Permanent stroke - No. (%) | 87 (1.9) | 10 (0.4) | 16 (1.3) | 20 (3.8) | 22 (6.3) | 19 (9.1) |
| >24hr ventilation - No. (%) | 458 (9.8) | 32 (1.3) | 59 (4.8) | 89 (16.9) | 122 (35.2) | 156 (75) |
| Reoperation - No. (%) | 293 (6.2) | 46 (1.9) | 42 (3.5) | 36 (6.8) | 67 (19.3) | 102 (49) |

**1.3 Autocorrelation and cross-correlation of test results for cardiac surgery patients.
eFigures 2-3** present autocorrelations and cross-correlations for the 8 biomarkers used for clustering, and autocorrelations for consecutive changes in these markers. Autocorrelations and cross-correlations were calculated between consecutive days (and between pre- and post-operation) and averaged over the patient cohort. All markers exhibit high autocorrelation over consecutive days, with four markers (RDW, PLT, BUN, CRE) having correlations continually above 0.9, reflecting slower dynamics than the other four markers. Three of the markers (WBC, ANION, GLU) also exhibit a type of ‘memory’, where correlation of day 5 values with pre-operative values is higher than correlations in the preceding days. This pattern may reflect a homeostatic memory, whereby patients return to their baseline. Four markers (RDW, PLT, BUN CRE) show high correlations between changes over consecutive days, reflecting a high momentum for these markers. In **eFigure 3**, changes in most biomarkers do not strongly correlate with each other. The key exceptions are blood cell populations (i.e. WBC x HCT, WBC x PLT and HCT x PLT), and renal indices (BUN x CRE), suggesting potentially significantly co-regulation of blood cell populations within this cohort.

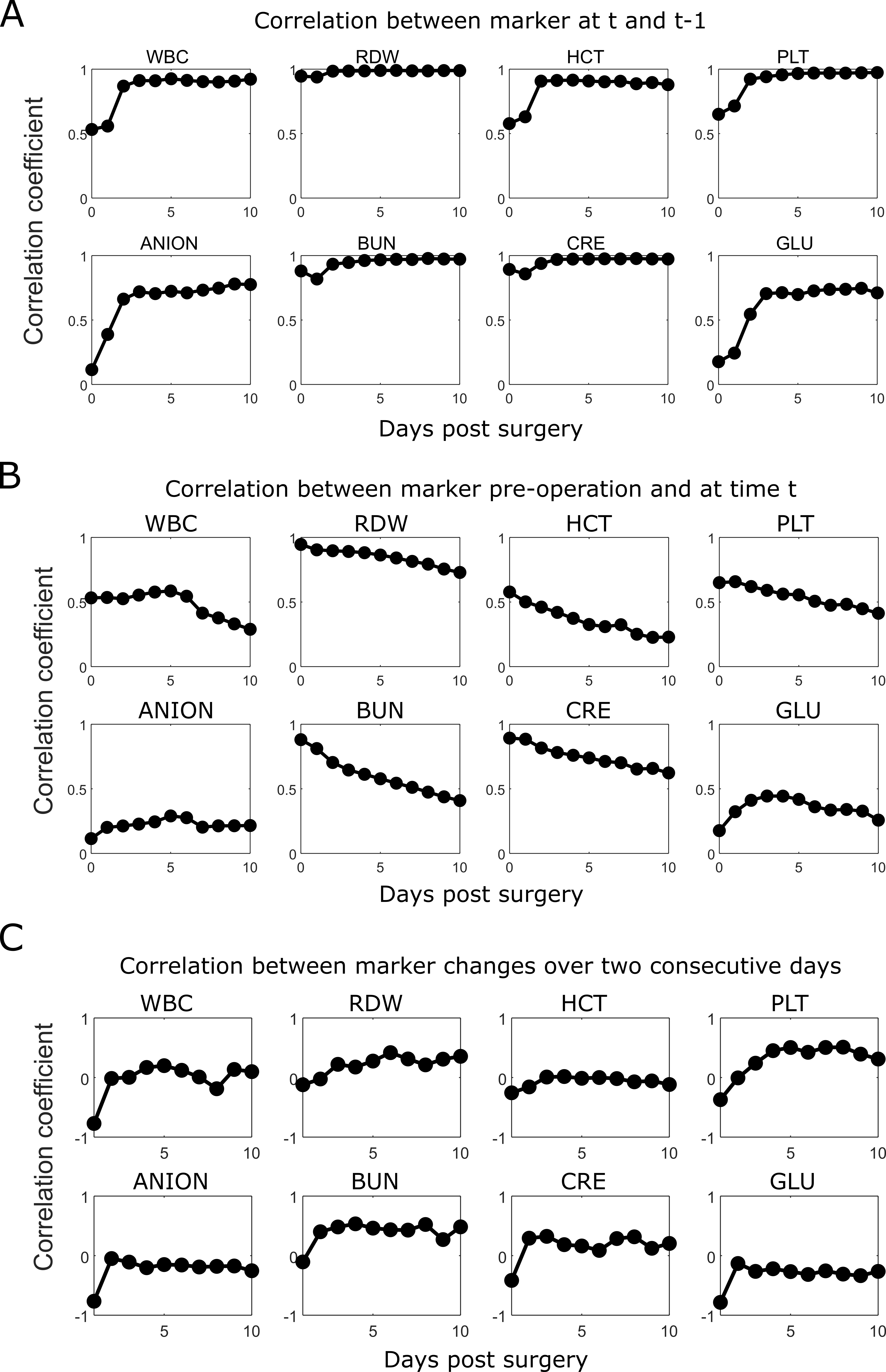

**eFigure 2 – Autocorrelations for test results throughout recovery from cardiac surgery**. Correlation coefficients are given for 8 tests (WBC, RDW, HCT, PLT, ANION, BUN, CRE, GLU) throughout recovery from cardiac surgery. Correlations are provided between values over consecutive days (A), between current and baseline values (B) and between marker changes over consecutive days (C).

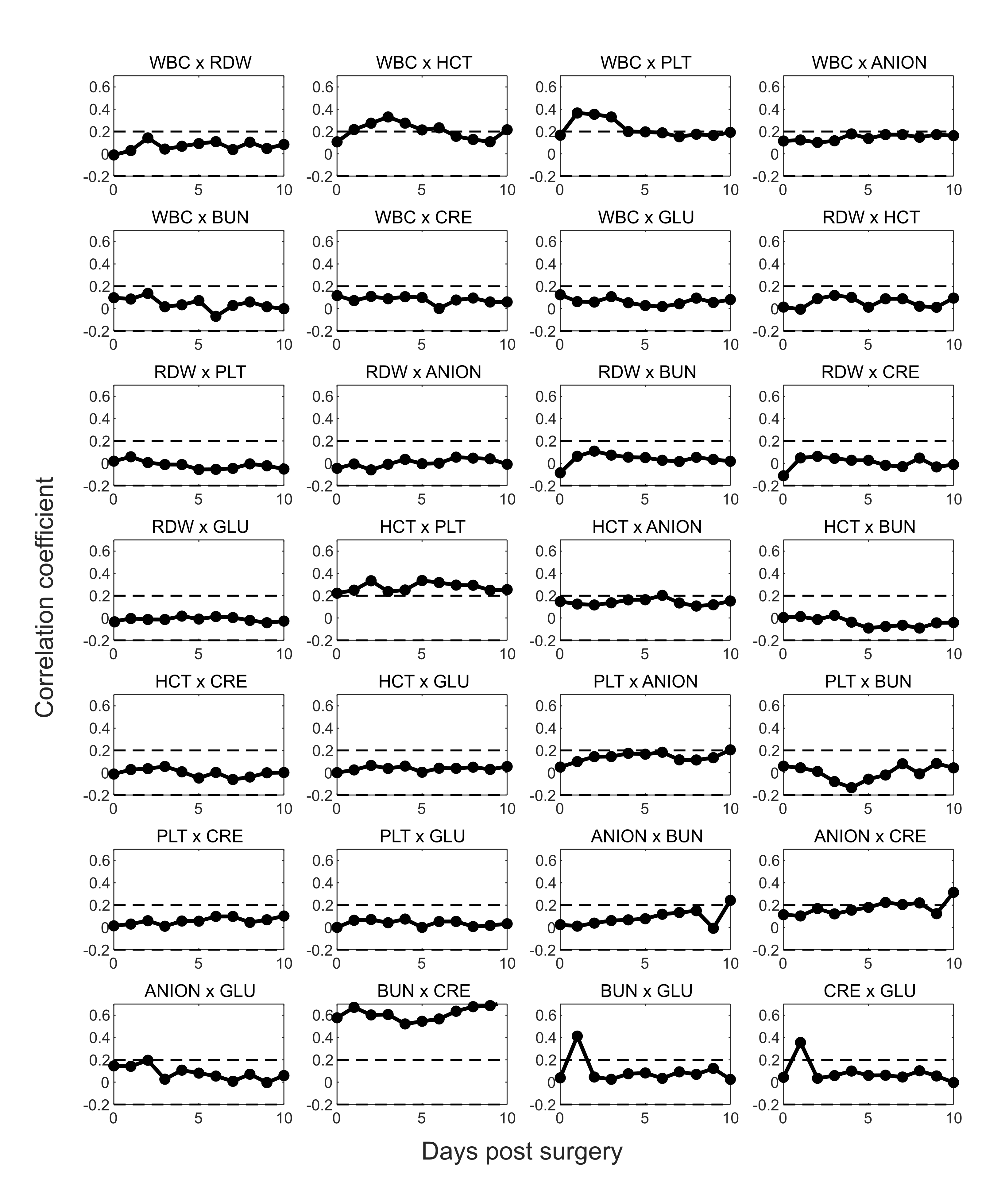
**eFigure 3 – Cross-correlations for biomarkers throughout recovery from cardiac surgery**. Cross-correlation coefficients are given for daily changes in 8 biomarkers (WBC, RDW, HCT, PLT, ANION, BUN, CRE, GLU) throughout recovery from cardiac surgery. Coefficients are the correlations between the change in each pair of biomarkers over the proceeding 24hr period. Most biomarker pairs show low cross-correlation, except for blood cell populations (WBC x HCT, WBC x PLT, HCT x PLT) and renal indices (BUN x CRE), suggesting strong coregulation of blood cell populations.

**1.4 Comparison of phase-plane analysis for other biomarker combinations**

The main manuscript described WBC-PLT trajectory analysis as a clinical risk stratification tool. This 2D distillation of the 8-biomarker set was chosen due to strong associations of WBC-PLT with mortality in the cardiac surgery cohort, and due to clinical insight, with WBC and PLT illustrating different dynamic speeds, and having clear clinical joint interpretability in the context of inflammation and homeostasis. While analysis of WBC-PLT dynamics provided strong stratification of favourable and unfavourable outcomes, this behaviour is not unique to these measures. We performed a parameter sweep (in the exploratory cohort only) using every combination of the 8 major biomarkers, comparing relative risk ratios at day 4 (as in **Fig 3**) between cardiac surgery patients with high (both > 80^th^) and low (both < 50^th^) position and direction percentiles. Through this sweep, WBC-PLT maintained the highest relative risk ratio (29.4: 1.7% to 52%) of all combinations. However, multiple other marker combination showed prognostic value, with the next best 5 combinations being WBC-RDW (relative risk 29.3: 1.8% to 54%), WBC-BUN (relative risk 21.5: 2.3% to 49%), and PLT-ANION (relative risk 18.2: 4% to 69%). A full list of parameter combinations and relative risks is given in **eTable 3**. In **eFigure 3** we present the overall mean trajectory, and trajectories for each cluster using those 3 alternate pairings. The adverse outcome likelihood and average increased length of stay stratified by day 4 position and direction percentiles are given in **eTable 3**.

As can be seen in **eTable 3-4** and **eFigure 3**, while other biomarker combinations generate similarly high relative risks there is overall less clear dominance of a single trajectory as seen for WBC-PLT, with the potential exception of PLT x ANION. Equally, considering results in **eTable 3-4**, while the relative risks of these other combinations are quite high, the number of patients with >80^th^ position and direction percentile is significantly smaller, reflecting lower correlation between negative position and direction, and lower sensitivity for identifying negative outcomes. Finally, there is also significant advantage to combinations such as WBC x PLT that require a single test, compared to biomarker combinations which involve both the CBC and BMP. Using pairs from the same test avoids any potential issues related to misalignment of the times at which the test samples are collected and analysed, which may be important when analysing fast changing dynamics (i.e. particularly during the first few days of recovery).

**eTable 3 – Adverse outcome stratification by day 4 position and direction percentiles for various biomarker pair combinations in the cardiac surgery cohort**

|  |  | Adverse outcome likelihood | |  |
| --- | --- | --- | --- | --- |
| Marker 1 | Marker 2 | Position and direction < 50th percentile | Position and direction > 80th percentile | Relative risk |
| PLT | WBC | 1.8% | 51.6% | 29.4 |
| RDW | WBC | 1.8% | 53.8% | 29.3 |
| WBC | BUN | 2.3% | 49.4% | 21.5 |
| PLT | ANION | 3.8% | 69.0% | 18.3 |
| PLT | BUN | 3.1% | 55.5% | 18.2 |
| WBC | GLU | 2.1% | 33.7% | 16.3 |
| WBC | ANION | 3.3% | 52.0% | 15.9 |
| PLT | GLU | 3.5% | 50.4% | 14.3 |
| HCT | WBC | 3.5% | 48.3% | 13.6 |
| WBC | CRE | 3.6% | 47.3% | 13.3 |
| PLT | RDW | 3.5% | 43.7% | 12.4 |
| HCT | PLT | 4.0% | 48.7% | 12.1 |
| HCT | RDW | 2.2% | 26.7% | 12.1 |
| ANION | GLU | 5.5% | 47.4% | 8.7 |
| PLT | CRE | 5.3% | 46.3% | 8.7 |
| RDW | GLU | 4.2% | 25.0% | 5.9 |
| ANION | CRE | 9.0% | 50.0% | 5.5 |
| HCT | ANION | 5.3% | 27.3% | 5.2 |
| ANION | BUN | 4.8% | 21.7% | 4.5 |
| RDW | ANION | 5.1% | 17.6% | 3.5 |
| BUN | GLU | 4.9% | 16.7% | 3.4 |
| HCT | GLU | 8.1% | 25.8% | 3.2 |
| BUN | CRE | 6.6% | 18.8% | 2.8 |
| RDW | CRE | 7.4% | 20.0% | 2.7 |
| CRE | GLU | 10.8% | 24.8% | 2.3 |
| HCT | BUN | 3.7% | 7.4% | 2.0 |
| RDW | BUN | 2.3% | 4.5% | 1.9 |
| HCT | CRE | 10.4% | 7.7% | 0.7 |

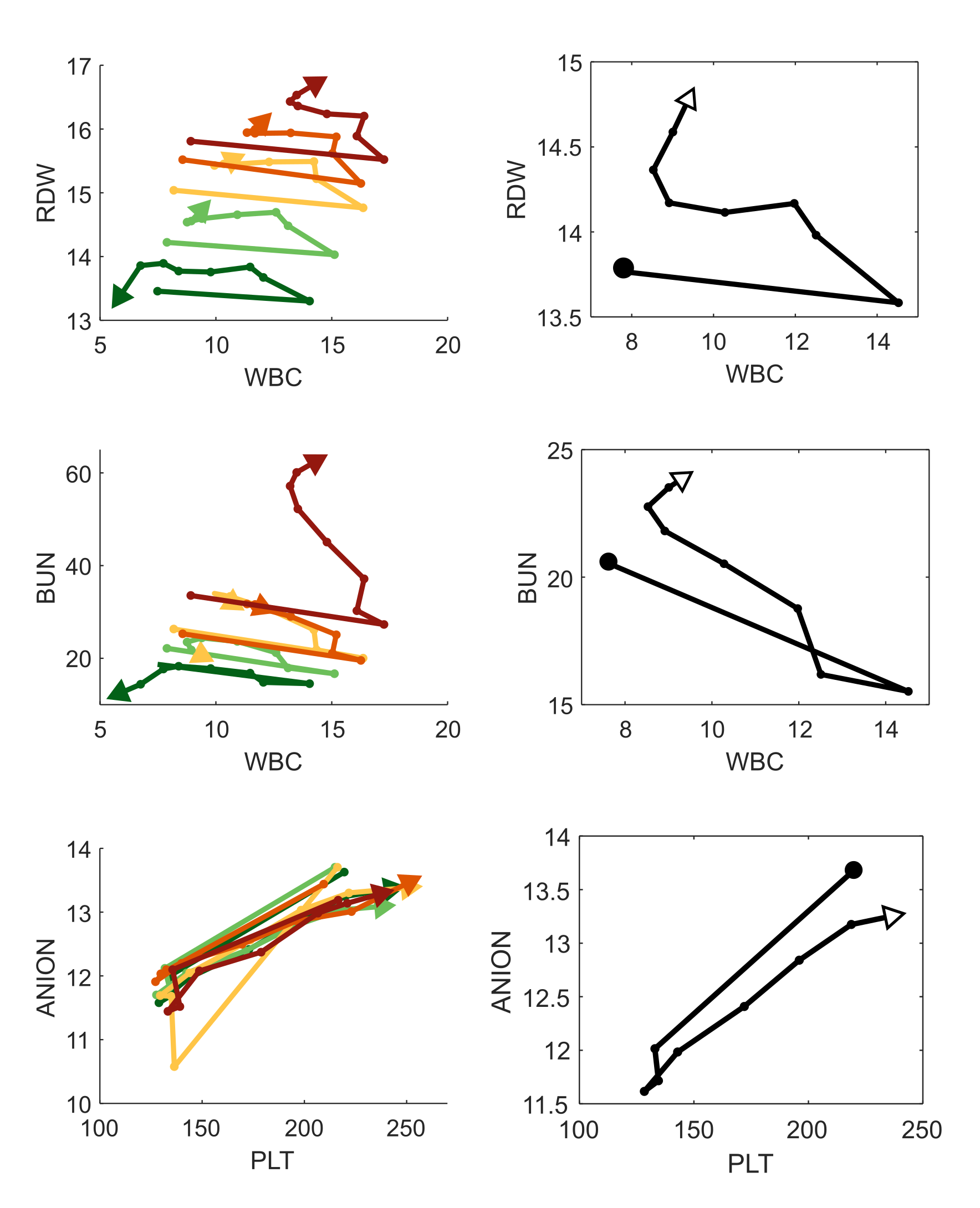
 **eFigure 3 – Cardiac surgery recovery trajectories for alternate biomarker pairs.** Mean trajectories for each cluster (left) and for patients with good outcomes (right) are given for three alternate pairs of biomarker combinations: WBC x RDW, WBC x BUN, and PLT x ANION. Each trajectory is from preop to day 7, with spacing between dots equal to 1 day.

**eTable 4 – Outcomes stratified by day 4 position and direction percentiles for alternate biomarker combinations in the cardiac surgery cohort**

|  |  | WBC x PLT | | | | | | | | |
| --- | --- | --- | --- | --- | --- | --- | --- | --- | --- | --- |
|  |  | Adverse outcomes likelihood | | | Remaining length of stay | | | Number of patients | | |
|  |  | Distance percentile | | | Distance percentile | | | Distance percentile | | |
|  |  | <50th | 50-80th | >80th | <50th | 50-80th | >80th | <50th | 50-80th | >80th |
| Position percentile | <50th | 1.75% | 3.3% | 14.7% | 2.1 | 3.0 | 5.9 | 399 | 211 | 68 |
|  | 50-80th | 6.8% | 7.4% | 30.6% | 3.5 | 5.2 | 10.7 | 162 | 122 | 72 |
|  | >80th | 25.0% | 23.0% | 52.6% | 7.2 | 9.2 | 15.1 | 56 | 61 | 133 |
|  |  | WBC x RDW | | | | | | | | |
|  |  | Adverse outcomes likelihood | | | Remaining length of stay | | | Number of patients | | |
|  |  | Distance percentile | | | Distance percentile | | | Distance percentile | | |
|  |  | <50th | 50-80th | >80th | <50th | 50-80th | >80th | <50th | 50-80th | >80th |
| Position percentile | <50th | 1.8% | 1.3% | 5.5% | 2.3 | 2.3 | 4.1 | 327 | 223 | 109 |
|  | 50-80th | 9.7% | 17.2% | 24.5% | 5.8 | 6.0 | 8.3 | 145 | 128 | 94 |
|  | >80th | 27.7% | 23.5% | 53.8% | 8.8 | 8.7 | 15.0 | 101 | 68 | 80 |
|  |  | WBC x BUN | | | | | | | | |
|  |  | Adverse outcomes likelihood | | | Remaining length of stay | | | Number of patients | | |
|  |  | Distance percentile | | | Distance percentile | | | Distance percentile | | |
|  |  | <50th | 50-80th | >80th | <50th | 50-80th | >80th | <50th | 50-80th | >80th |
| Position percentile | <50th | 2.3% | 2.0% | 6.0% | 2.3 | 3.1 | 3.3 | 348 | 204 | 117 |
|  | 50-80th | 8.3% | 14.3% | 29.6% | 4.8 | 4.7 | 9.4 | 157 | 91 | 81 |
|  | >80th | 21.9% | 31.7% | 49.4% | 8.4 | 9.8 | 15.5 | 105 | 82 | 85 |
|  |  | PLT x ANION | | | | | | | | |
|  |  | Adverse outcomes likelihood | | | Remaining length of stay | | | Number of patients | | |
|  |  | Distance percentile | | | Distance percentile | | | Distance percentile | | |
|  |  | <50th | 50-80th | >80th | <50th | 50-80th | >80th | <50th | 50-80th | >80th |
| Position percentile | <50th | 3.8% | 8.6% | 10.5% | 2.8 | 4.0 | 5.9 | 531 | 175 | 57 |
|  | 50-80th | 12.2% | 11.0% | 21.4% | 4.9 | 5.8 | 8.3 | 164 | 100 | 56 |
|  | >80th | 15.7% | 28.4% | 69.0% | 6.5 | 10.9 | 19.0 | 51 | 67 | 71 |

**1.5 Mean cardiac surgery WBC-PLT trajectories for patients stratified by gender, year, and STS risk cohort.**

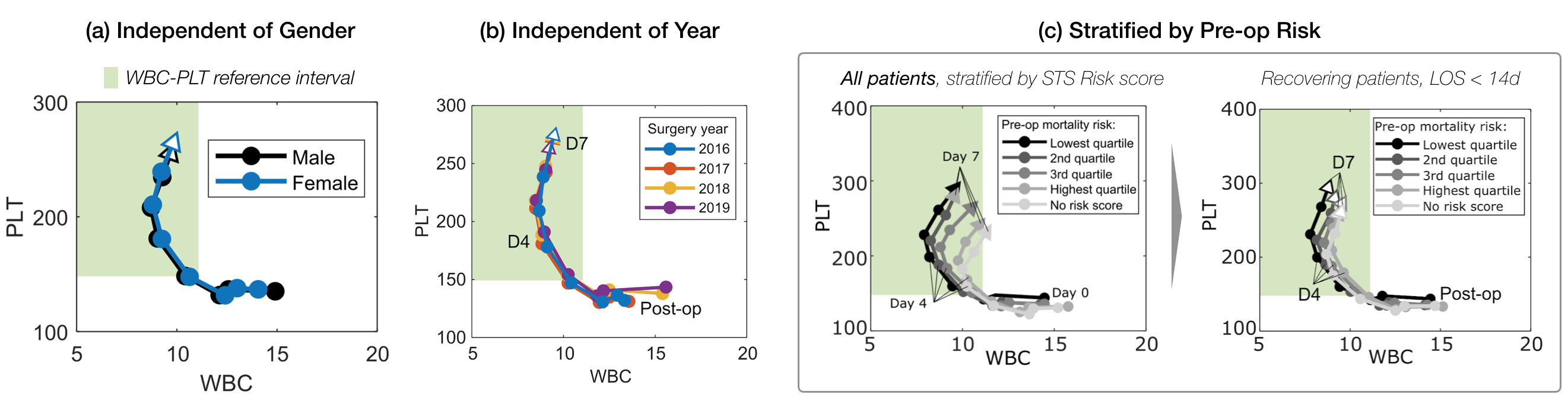
Fig 1 presents evidence for the invariance of the mean WBC-PLT response trajectory for cardiac surgery patients with favourable outcomes, independent of stratification by a wide range of factors. For completeness, in eFigure 4 we illustrate these mean trajectories when stratified by additional clinical and historical parameters, as in Fig 1C. (a) The mean trajectory for patients with favourable outcomes is not significantly different for males or females; (b) nor when stratified by the year of surgery. (c) When including all patients, including those with adverse outcomes, stratification by STS mortality risk identifies distinct WBC-PLT phase plane trajectories as a function of risk quartile; for recovering patients (survivors, LOS < 14d) however, trajectories are well-aligned.

**eFigure 4 – Mean WBC-PLT trajectories for cardiac surgery patients stratified by gender, year, and STS risk cohort.**

**1.6 Mean cardiac surgery WBC-PLT trajectories for patients with unfavourable outcomes.**

**Fig 1** presents evidence for the invariance of the mean WBC-PLT cardiac surgery response trajectory for patients with favourable outcomes, independent of stratification by a wide range of factors (age, surgery type, hospital, pre-op values, etc.). **eFigure 5** illustrates the equivalent trajectories for patients within three groups of unfavourable outcomes: surviving patients with LOS of 14-21 days, surviving patients with LOS of 21+ days, and patients who did not survive. Unlike the various categorisations of the favourable outcome cohort, the mean WBC-PLT trajectories for patients with unfavourable outcomes vary significantly and show a much lower coherence with each other. This provides further evidence that there is one dominant archetype of favourable inflammatory response, while there are many different archetypes of an unfavourable inflammatory response.

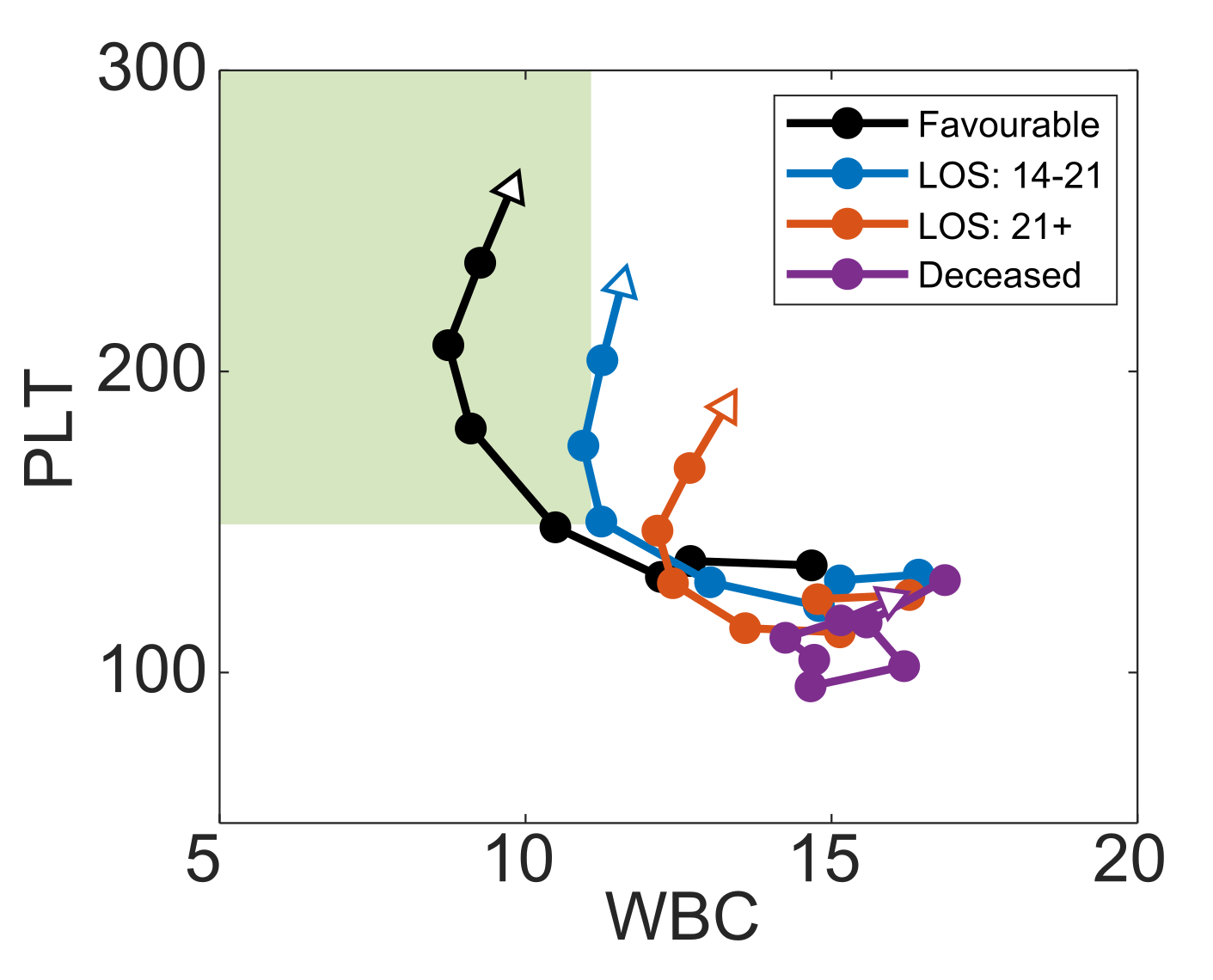

**eFigure 5 – Mean WBC-PLT trajectories for cardiac surgery patients with unfavourable outcomes.** Mean trajectories are given for patients who survive with post-op hospital stays of 2-3 weeks, greater than 3 weeks, and for patients who do not survive. For comparison, the reference trajectory (**Fig 1C**) is also included. Unlike the results in **Fig 1C**, there is much lower coherence between the mean trajectories of patients with different kinds of unfavourable outcomes.

**1.7 Example WBC-PLT trajectories across the cohort**

**Fig. 1** illustrates the mean WBC-PLT recovery trajectory for patients who underwent cardiac surgery. While this mean response is robust to a wide variety of factors, actual patient trajectories will not perfectly adhere to this trajectory. To illustrate the degree of patient variation in the MGH cardiac surgery cohort, in **eFigure 6** we present patient trajectories from post-op days 1 to 7. Trajectories of the 5 patients closest to the 0^th^, 25^th^, 50^th^, and 75^th^ percentiles for mean deviation over days 1-7 were plotted. As the figure shows, patients below the 50^th^ percentile have close adherence to the mean trajectory, while patients at the 75^th^ percentile deviate much more significantly, and with high variance. These results align with the primary finding of a dominant healthy recovery trajectory shape that the typical patient follows. For the subset of patients who deviate from this curve, there is little consistency in trajectories. It should be noted that **eFigure 6** trajectories were only chosen from patients who stayed at least 7 days in hospital, and may be biased towards adverse trajectories (given ~50% of patients discharge before day 7).

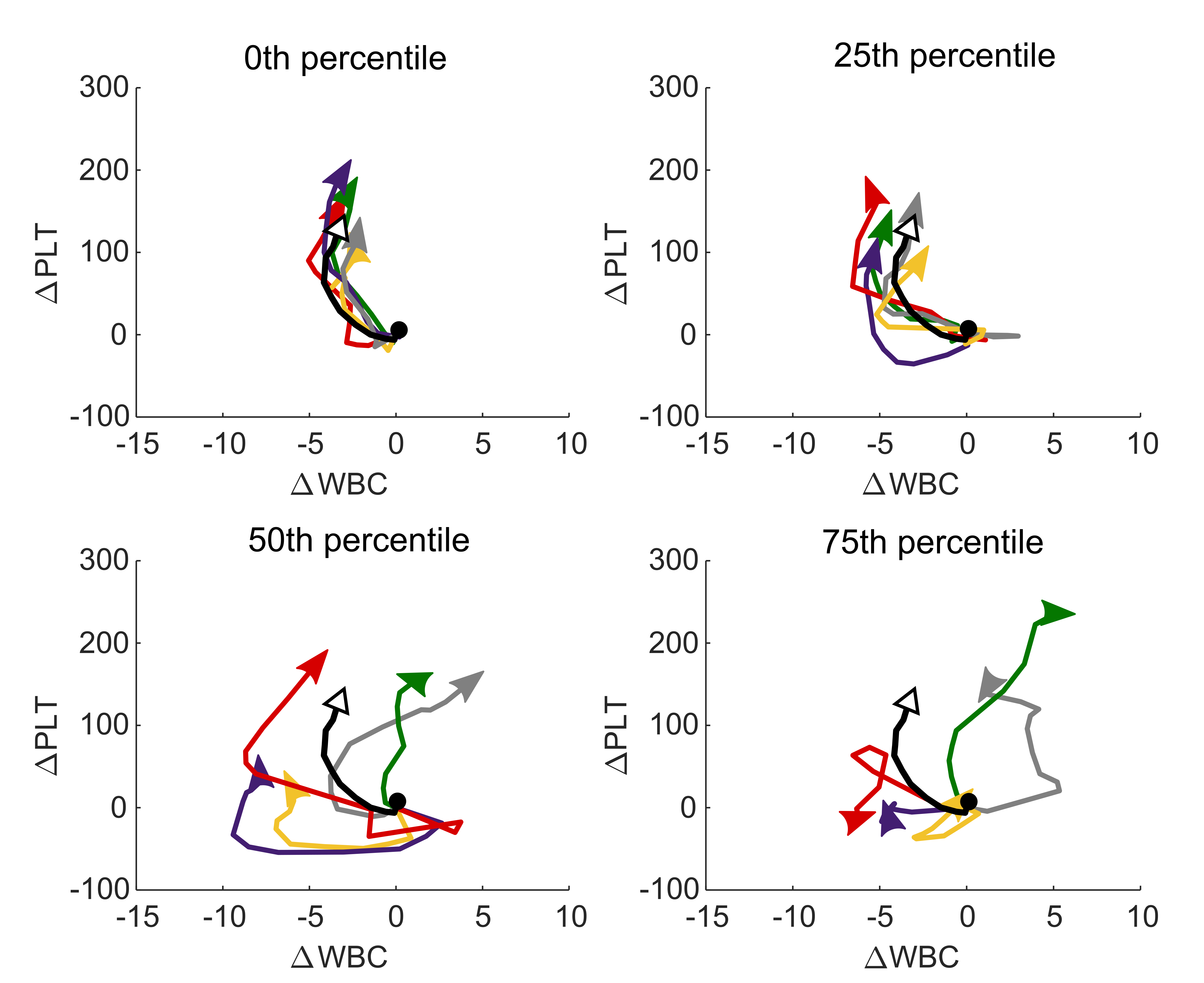

**eFigure 6 – Example cardiac surgery WBC-PLT trajectories stratified by average degree of deviation.** The 5 patient trajectories closest to the 0^th^, 25^th^, 50^th^ and 75^th^ percentile of average deviation (from day 1 to day 7) from the mean WBC-PLT trajectory are given, from post op day 1 to day 7. The 0^th^ and 25^th^ percentile trajectories adhere closely to the mean trajectory. The 50^th^ percentile exhibits high variance early on, but eventually adheres to the shape of the mean trajectory. No consistent patterns are seen in the 75^th^ percentile trajectory. Patient selection was limited to MGH cohort patients with hospital stays > 7 days, and as such is biased towards adverse trajectories.

**1.8 Surgery trajectories stratified by length of hospital stay.**

**Fig 1C.ii** shows the mean WBC-PLT trajectory for the cardiac surgery cohort stratified by LOS. **eFigure 7** presents similar stratifications for the other 6 surgery cohorts. Unlike the cardiac surgery cohort, some of the other surgery cohorts show significant differences in starting position and WBC perturbation size when stratified by LOS. However, after normalization by post-op day 1 WBC and PLT, and the injury size (size of pre-op to post-op WBC increase), all 6 cohorts show similar mean trajectory shapes independent of LOS.

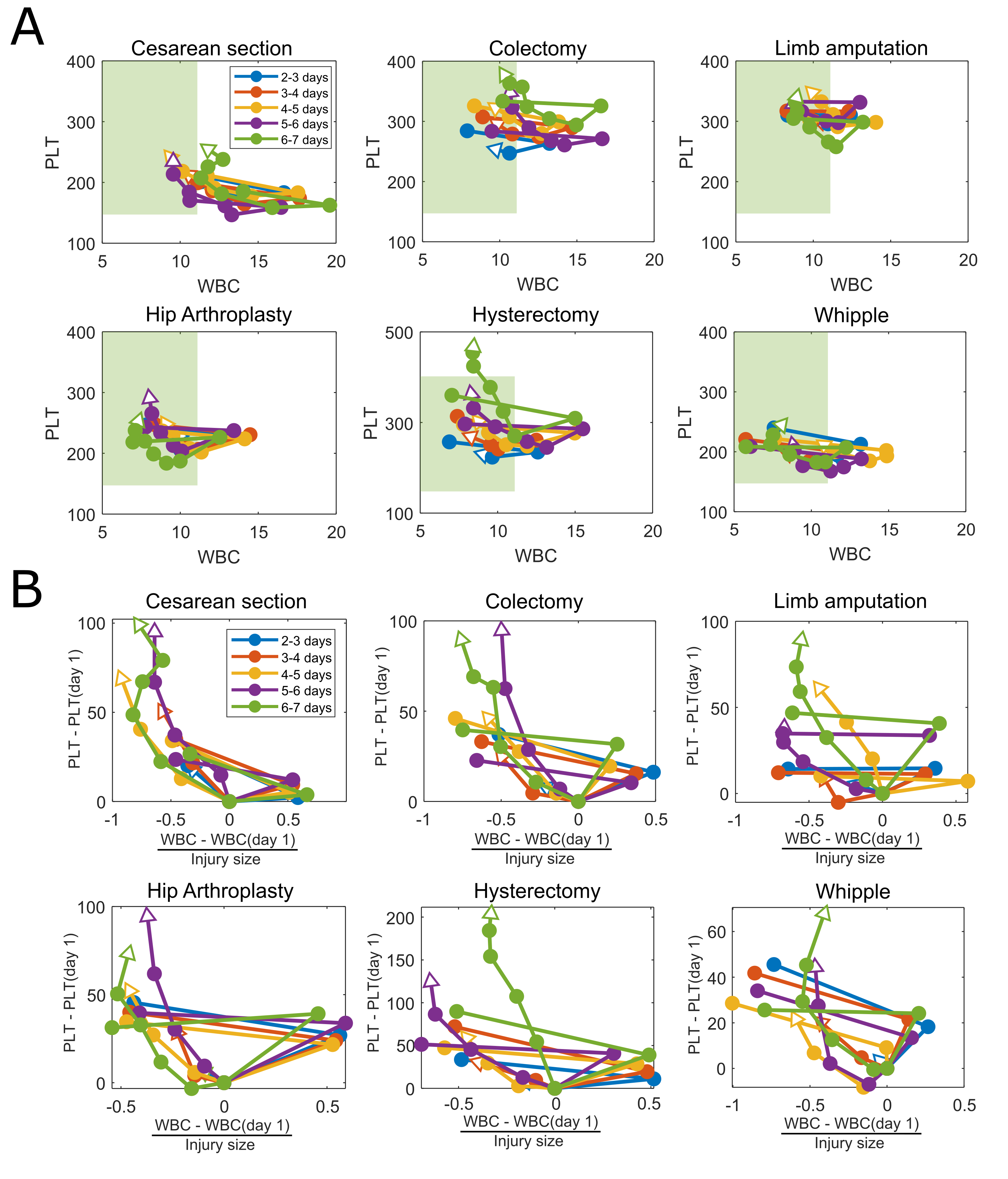

**eFigure 7 – Stratification of surgical cohort WBC-PLT trajectories by length of hospital stay (LOS).** Results are presented for patients with LOS of 2-3, 3-4, 4-5, 5-6, and 6-7 days, without normalization (A), and with normalization (B) based on post-op day 1 WBC and PLT, and injury size (post-op WBC – pre-op WBC). While patients with different LOS have different baseline WBC-PLT and injury size, the shape of the mean normalized trajectories are consistent.

**1.9 Model fits for other inflammatory cohorts**

**Fig 2** presents exponential and linear model fits for average WBC and PLT counts post alignment (full details described in main manuscript methods). **eFigure 8** presents the equivalent fits for the cohorts not included in **Fig 2**: Cesarean, colectomy, Whipple, stroke, and C. diff. colitis. As shown, each of the other inflammatory cohorts demonstrates the same average behaviour of an exponential decay in WBC and a lagged linear rise in PLT.

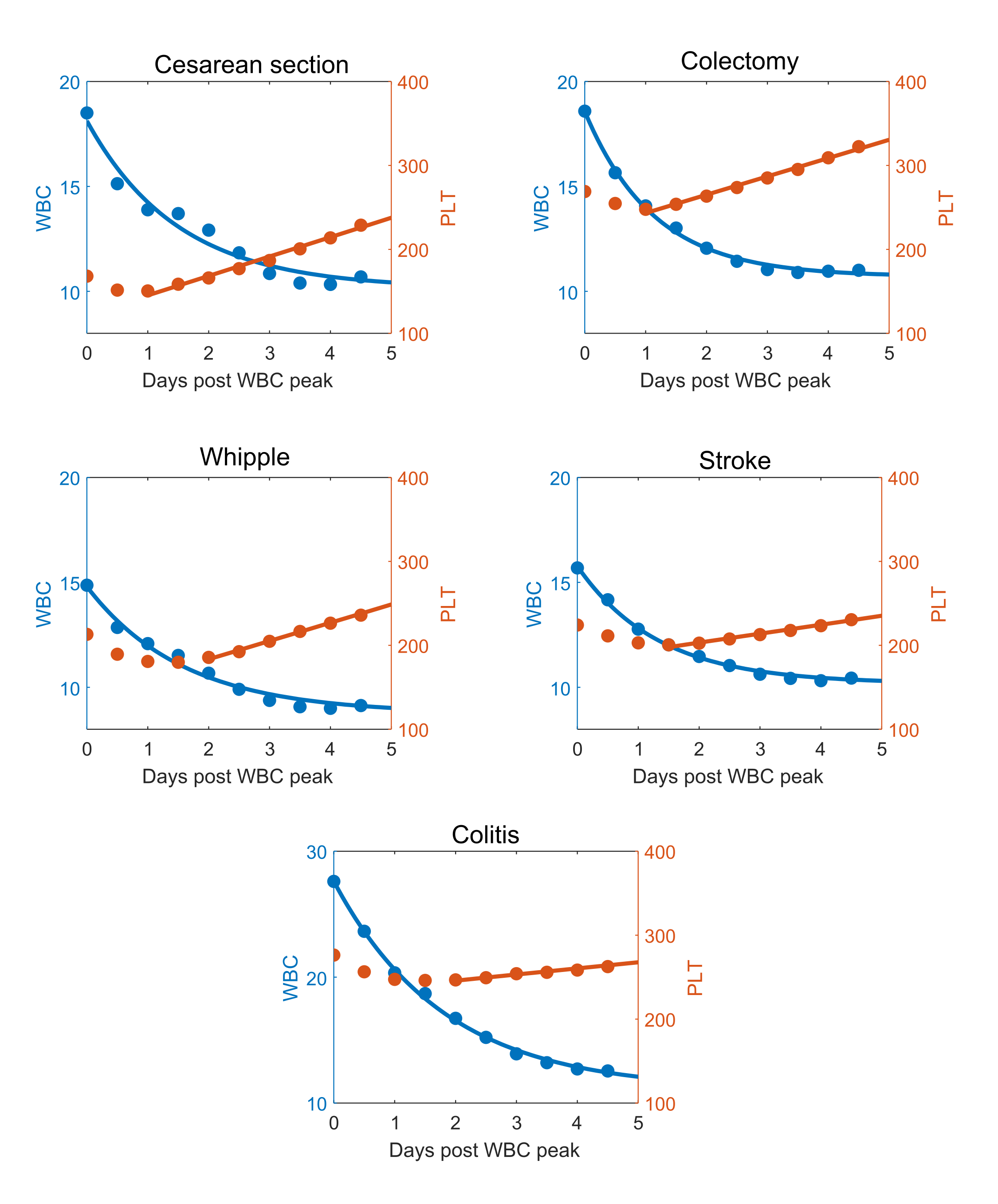

**eFigure 8 – Exponential and linear model fits for other inflammatory cohorts.** Mean WBC and PLT data for 5 days post WBC peak are given for 5 inflammatory cohorts: Cesarean, colectomy, Whipple, stroke, and C. diff. colitis. Fits of an exponential model (WBC) and linear model (PLT) are included.

**1.10 Stratifications of the COVID and myocardial infarction WBC-PLT trajectories by demographic and clinical factors.**

**Fig 1C** presents the mean WBC-PLT trajectory for the cardiac surgery cohort stratified by a range of demographic and clinical factors. **eFigure 9** shows similar stratifications for one of the ischemia cohorts (myocardial infarction) and one of the infection cohorts (COVID-19). In both cases, the shape of the mean WBC-PLT trajectory is conserved across stratifications by gender, age, LOS, and admission WBC and PLT counts.

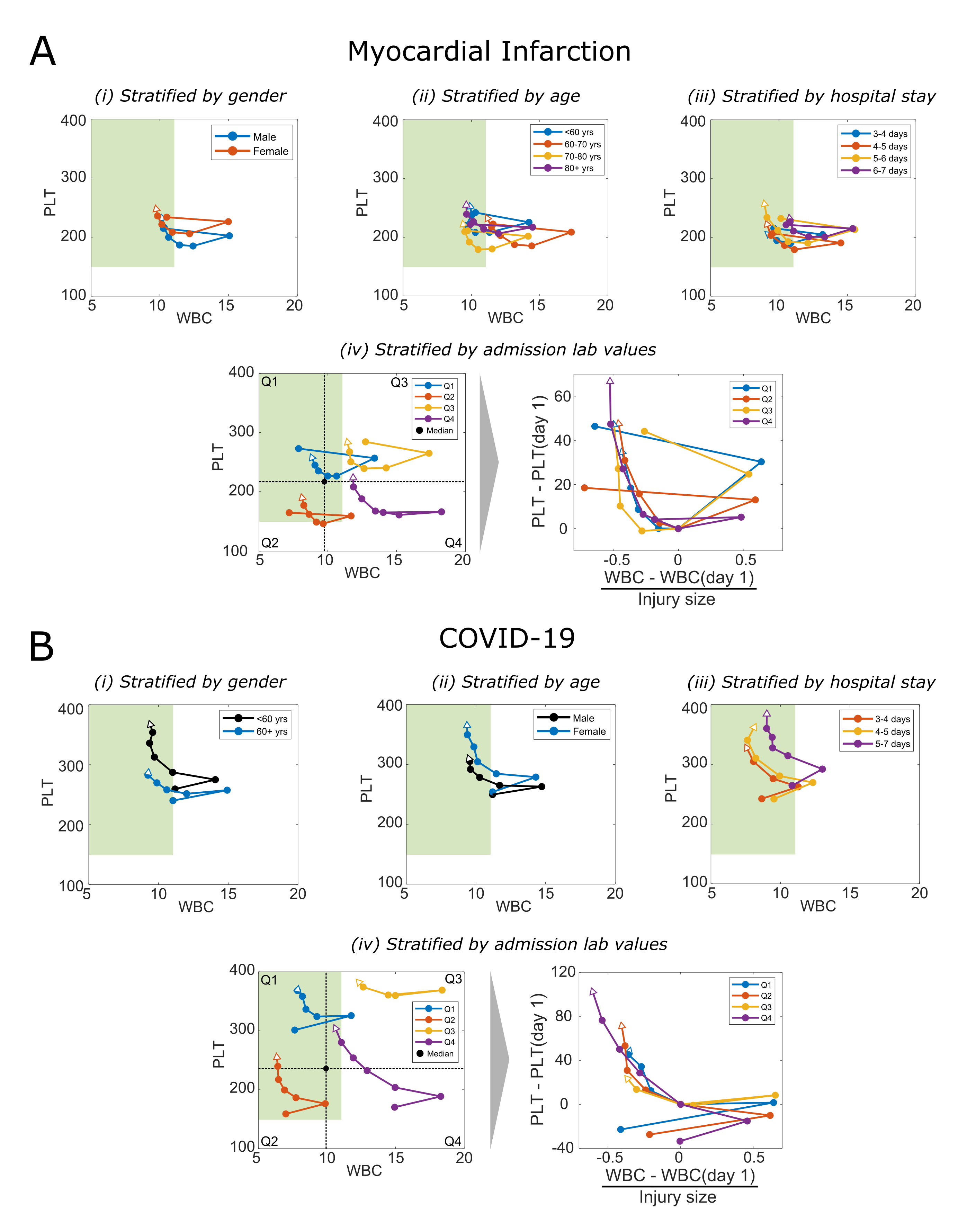

**eFigure 9 – Mean WBC-PLT trajectories stratified by demographic and clinical factors for myocardial infarction and COVID-19 cohorts.** Stratification by gender, age, and LOS is associated with admission WBC-PLT, but the shape of the mean trajectory is similar independent of gender, age, LOS, and admission WBC-PLT. Both COVID-19 and myocardial infarction cohorts have been aligned as described in the main text methods.

**1.11 Risk ratios for other inflammatory cohorts**

**Fig 3** of the main manuscript presents adverse outcome risks stratified by position and direction percentiles for 5 inflammatory cohorts. For completeness, in **eFigure 10** we present equivalent results for the remaining 6 cohorts: Amputation, Cesarean section, hysterectomy, Whipple, stroke, and C. diff. colitis. Note that due to sample size constraints, only results for stroke represent out-of-sample testing, where percentiles and thresholds were defined from the exploratory set, and outcome rates were calculated from the validation set. For the other five cohorts, percentiles, thresholds, and outcome rates were calculated from the overall dataset. The colitis dataset was too small to allow for stable estimates of outcome rates, even with in-sample testing. Results have been included for completeness but should be interpreted with caution. In all cases there are significant increases in risk for patients whose day 4 position and direction are above the 80^th^ percentile, compared to patients who are below the 50^th^ percentile for both.

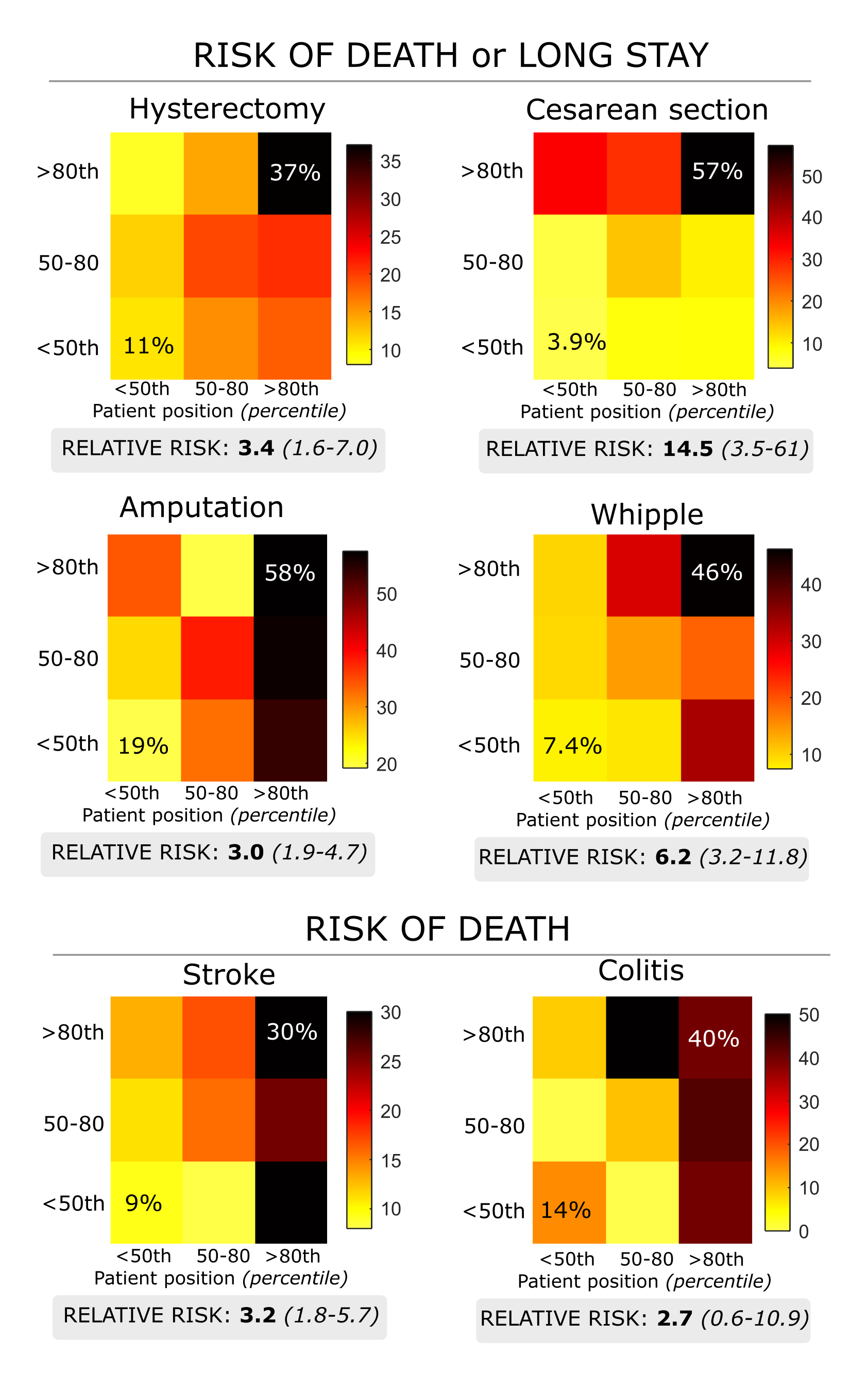

**eFigure 10 – Risk of adverse outcomes stratified by day 4 positional and directional risk.** In similar format to **Fig 3**, patient risk of death or long stay, stratified by day 4 positional and directional percentiles are given for the remaining 6 inflammation cohorts. In each case, patients with position and direction above the 80^th^ percentile have significantly elevated risk comparative to patients whose position and direction is below the 50^th^ percentile.

**1.12 Risk ratios for COVID-19 without cohort alignment**

**Fig 3** of the main manuscript presents mortality risk for the COVID-19 cohort, stratified by position and direction percentiles. To account for potential differences between timing of diagnosis and timing of peak inflammatory response, patients were aligned based on the timing of their maximum WBC count within the first 72hrs post admission. In **eFigure 11** we present the equivalent risk stratification in the COVID-19 cohort without alignment of WBC values. As shown, while the overall outcome prevalence rates differ from those in **Fig 3**, the magnitude of risk stratification is quantitatively similar.

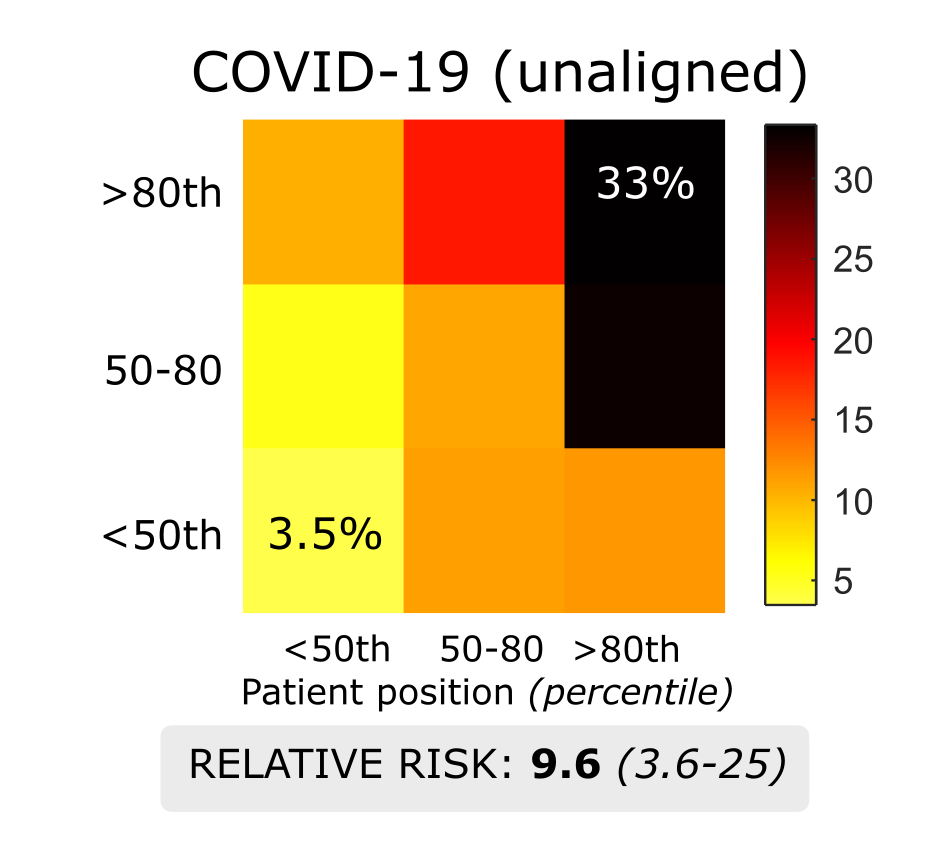

**eFigure 11 – COVID-19 mortality risk stratification by position and direction percentiles without alignment.** While the exact prevalence rates differ from the aligned results in **Fig 3**, the overall magnitude of risk stratification from patients with position and direction < 50^th^, to those with position and direction > 80^th^ is quantitatively similar.

**1.13 WBC-PLT trajectory analysis reference position and direction charts**

**Fig 3** shows how WBC-PLT trajectory analysis can provide risk stratification of acute inflammatory cohorts. This stratification was performed by comparison of patient position and direction to time-dependent reference charts. In **eFigures 12-16** we present the reference position and direction charts for post-op (post admission for non-surgical cohorts) days 1 to 6, for cardiac surgery, hip arthroplasty, colectomy, COVID-19 and myocardial infarction. We also present the corresponding likelihood of death (COVID-19, MI) or death/long stay (cardiac surgery, hip arthroplasty, colectomy) stratified by position and direction percentiles on each of those days in **eTable 5.**

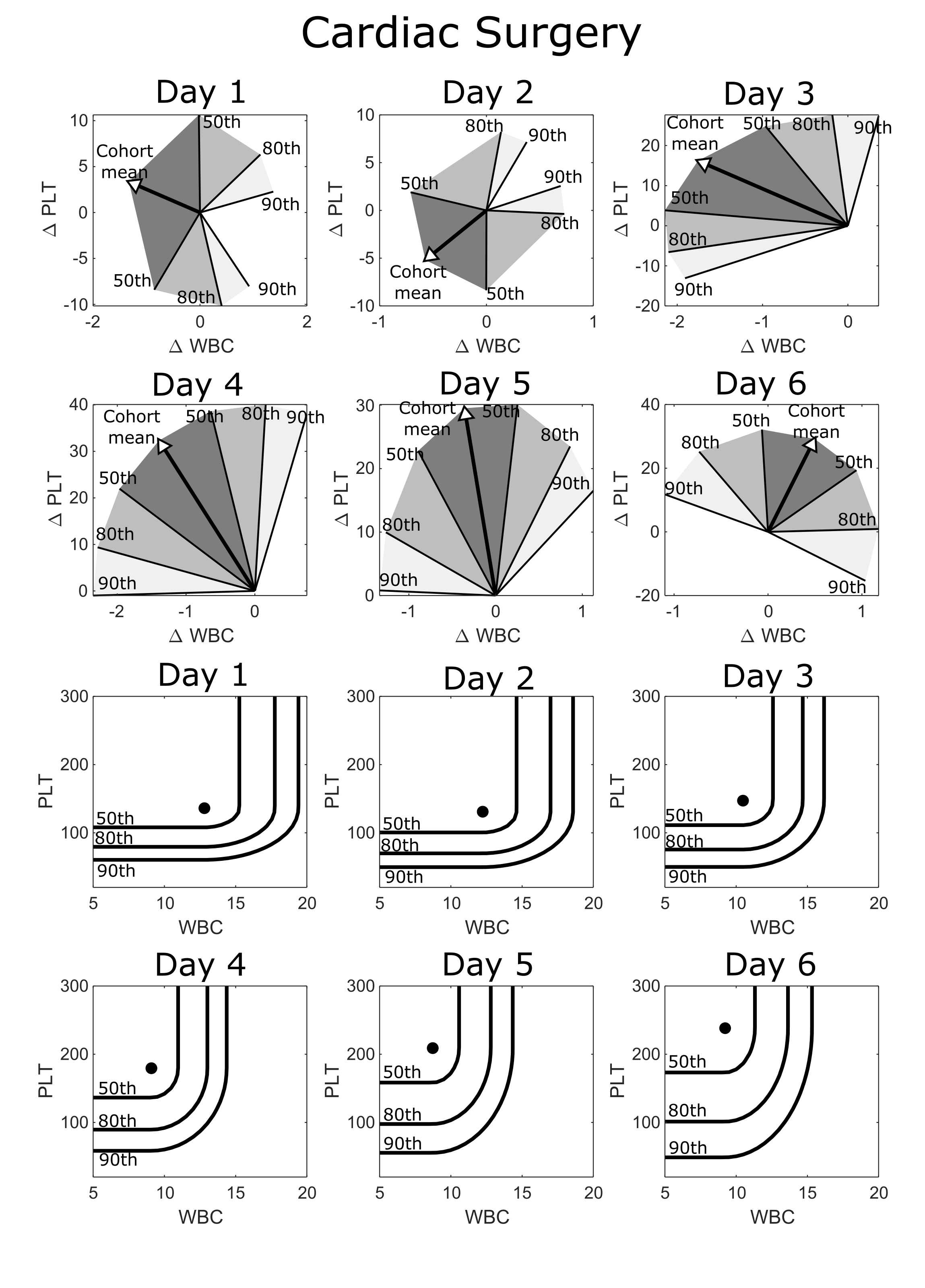

**eFigure 12 – Reference position and direction percentiles charts for post-op days 1 through 6 for cardiac surgery.**

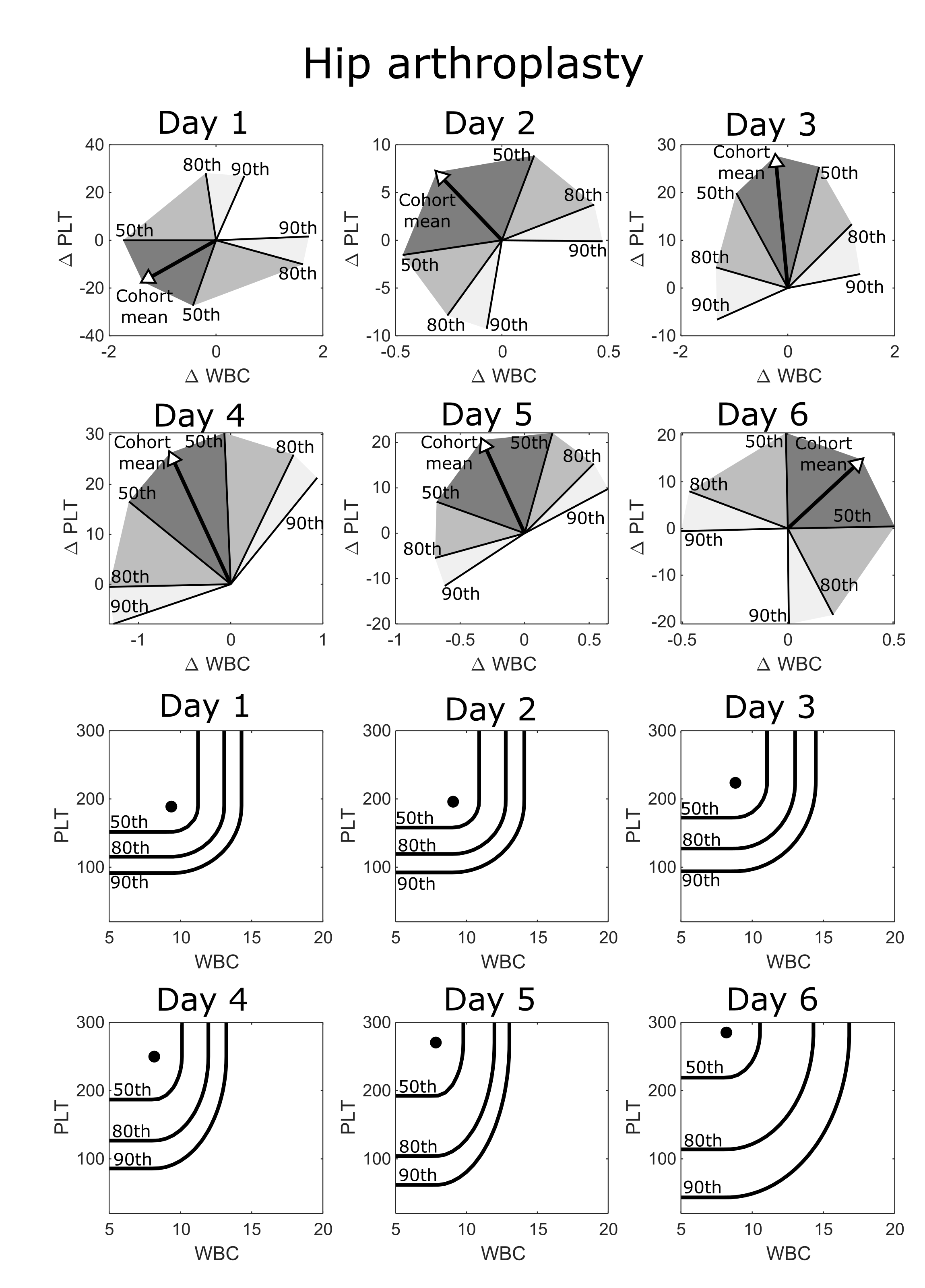

**eFigure 13 – Reference position and direction percentiles charts for post-op days 1 through 6 for hip arthroplasty.**

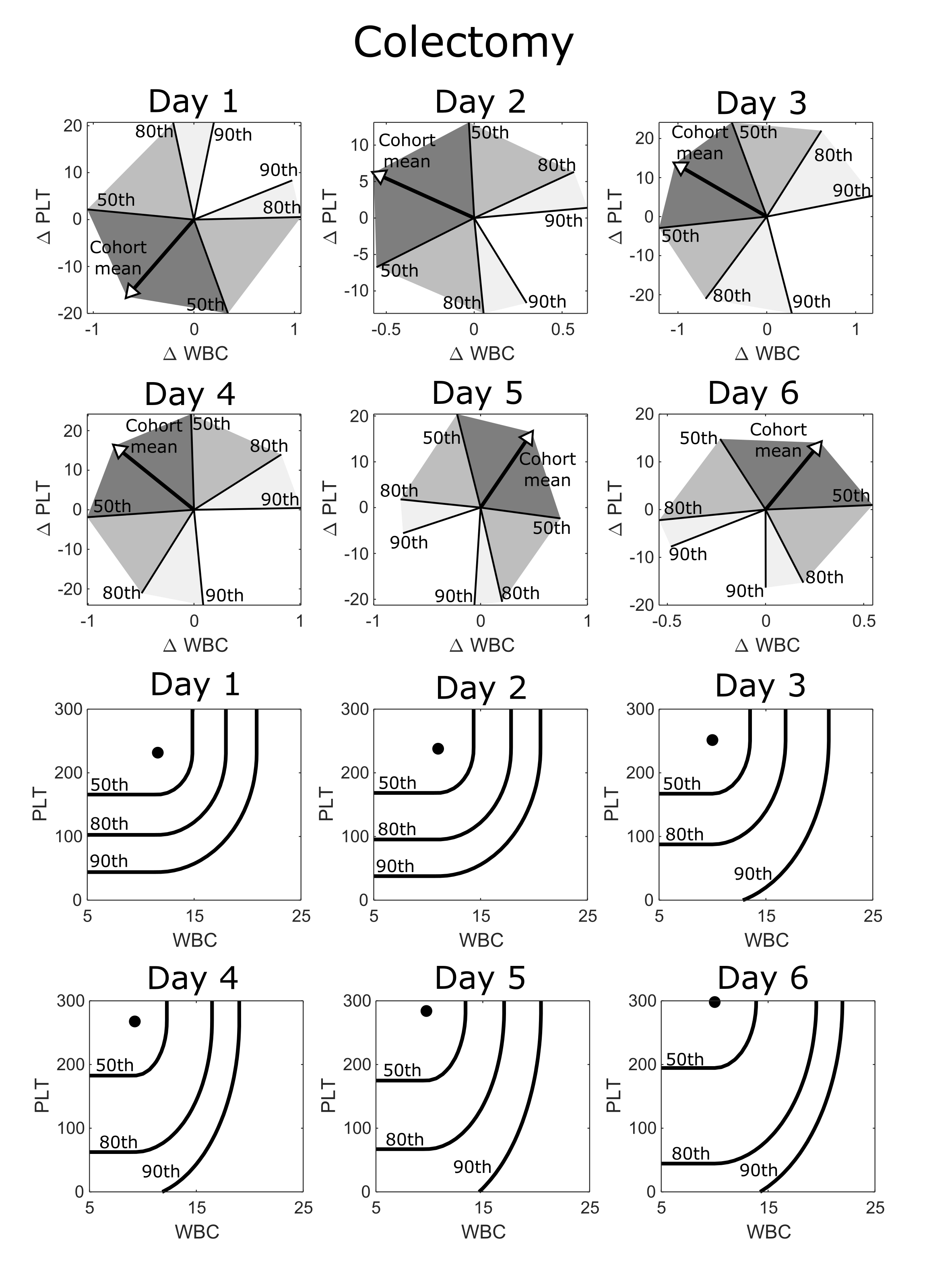

**eFigure 14 – Reference position and direction percentiles charts for post-op days 1 through 6 for colectomy.**

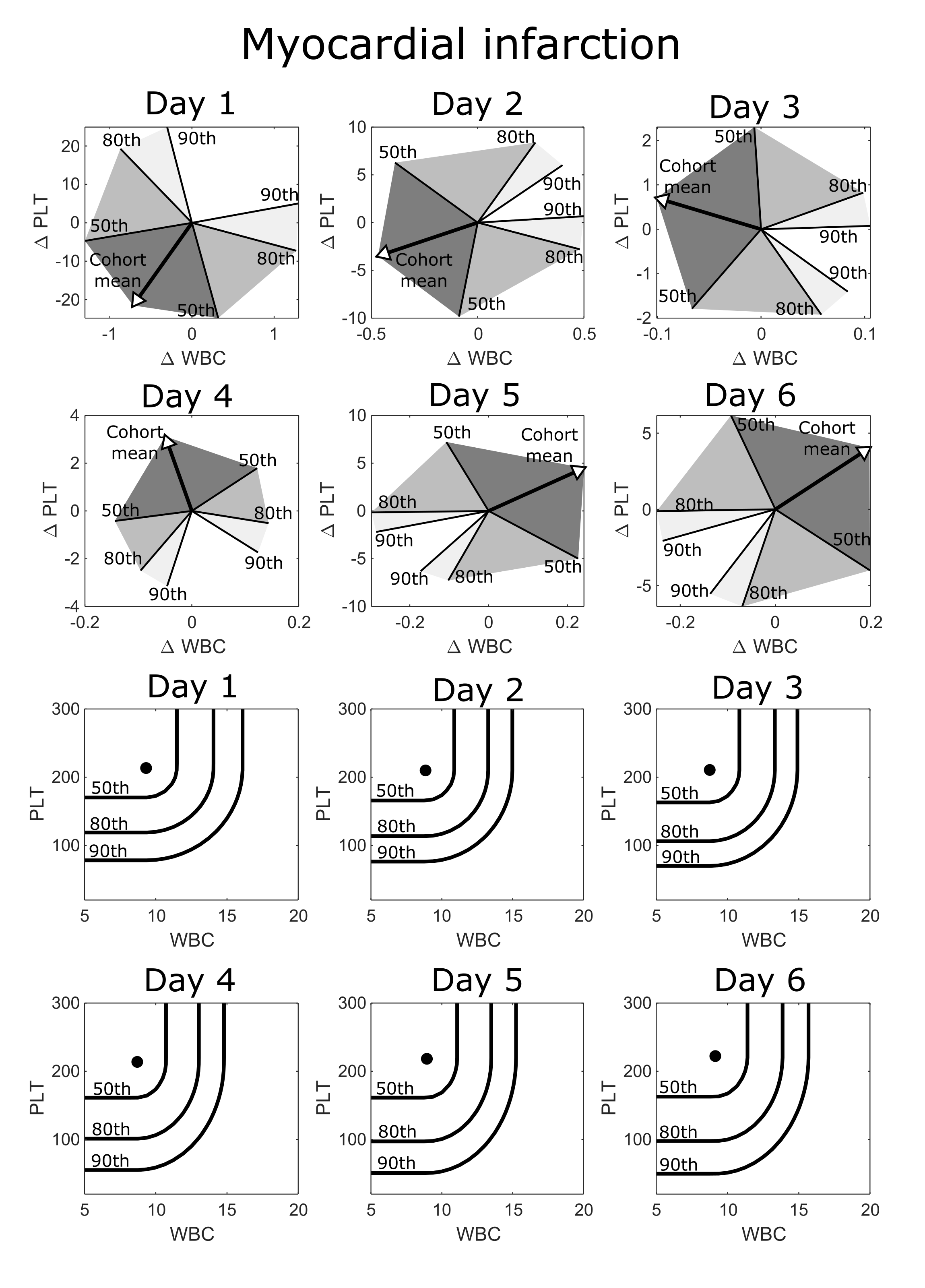

**eFigure 15 – Reference position and direction percentiles charts for post admission days 1 through 6 for myocardial infarction.**

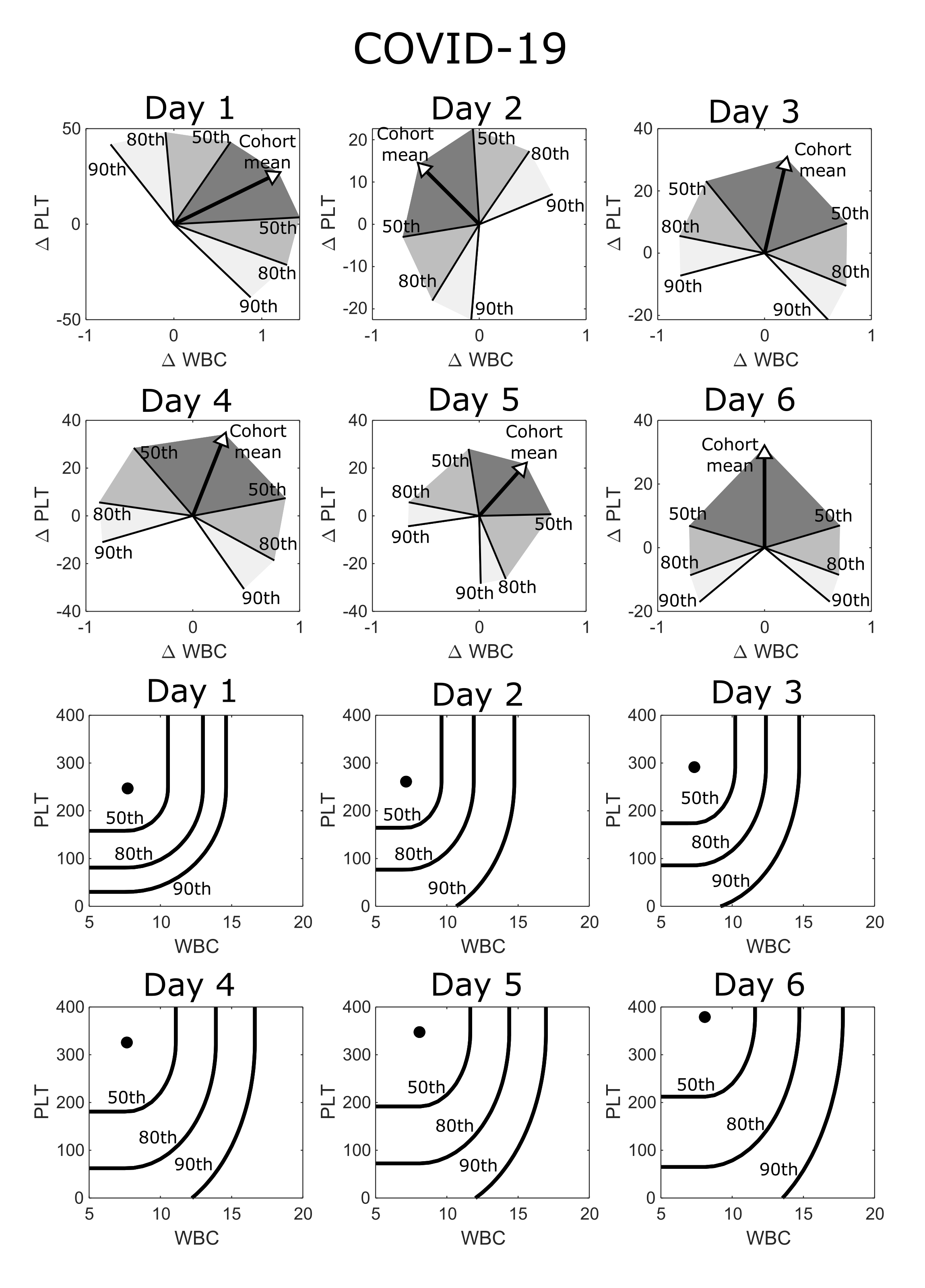

**eFigure 16 – Reference position and direction percentiles charts for post admission days 1 through 6 for COVID-19, post alignment using maximum WBC count during first 72hrs.**

|  |  |  | Cardiac Surgery | | | Hip arthroplasty | | | Colectomy | | | Myocardial infarction | | | COVID-19 | | |
| --- | --- | --- | --- | --- | --- | --- | --- | --- | --- | --- | --- | --- | --- | --- | --- | --- | --- |
|  |  |  | Mortality or LOS > 14 - % (Number in sub-group) | | | Mortality or LOS > 10 - % (Number in sub-group) | | | Mortality or LOS > 14 - % (Number in sub-group) | | | Mortality - % (Number in sub-group) | | | Mortality - % (Number in sub-group) | | |
|  |  |  | Distance percentile | | | Distance percentile | | | Distance percentile | | | Distance percentile | | | Distance percentile | | |
|  |  |  | <50th | 50-80 | >80th | <50th | 50-80 | >80th | <50th | 50-80 | >80th | <50th | 50-80 | >80th | <50th | 50-80 | >80th |
| Day 1 | Position percentile | <50th | 3.9% (571) | 8.6% (209) | 1.8% (56) | 1.6% (373) | 2% (248) | 5.8% (155) | 6.6% (211) | 10.2% (98) | 8% (88) | 5.2% (806) | 4.9% (408) | 5.6% (306) | 4.7% (106) | 5.6% (54) | 6.5% (31) |
|  |  | 50-80 | 9.7% (259) | 11.8% (93) | 12.2% (49) | 2.5% (241) | 4.8% (146) | 0% (81) | 11.4% (123) | 7.8% (51) | 16.7% (48) | 8% (439) | 9.1% (263) | 10.3% (175) | 7.8% (64) | 23.1% (13) | 12.5% (16) |
|  |  | >80th | 26.9% (134) | 25% (52) | 39.1% (64) | 14.6% (151) | 10% (120) | 6.3% (95) | 47.7% (65) | 33.3% (60) | 31% (42) | 18.8% (276) | 17.1% (234) | 21.3% (202) | 25% (32) | 26.1% (23) | 25% (16) |
| Day 2 | Position percentile | <50th | 3.5% (367) | 5.9% (236) | 3.3% (183) | 3% (302) | 6.1% (197) | 2.3% (86) | 7% (213) | 10.3% (107) | 15.6% (45) | 3.7% (758) | 7.1% (520) | 6% (250) | 0.9% (111) | 8.9% (45) | 10.5% (19) |
|  |  | 50-80 | 10.2% (215) | 15.9% (138) | 11.3% (71) | 2.2% (183) | 3.6% (138) | 4.7% (64) | 11.7% (103) | 11.3% (62) | 18.4% (38) | 9.3% (454) | 8.8% (228) | 7% (157) | 10.2% (49) | 17.4% (23) | 10% (20) |
|  |  | >80th | 22.3% (103) | 25.7% (101) | 31% (71) | 12.6% (87) | 12.1% (107) | 21.5% (65) | 38.1% (63) | 40.6% (32) | 47.1% (51) | 16.2% (370) | 18.6% (194) | 24.3% (181) | 22.7% (22) | 9.1% (11) | 52% (25) |
| Day 3 | Position percentile | <50th | 1.4% (370) | 1.9% (269) | 10.5% (143) | 6.5% (186) | 3.5% (86) | 19.4% (36) | 7.1% (184) | 5.4% (74) | 13.8% (58) | 6.2% (616) | 6.5% (401) | 4.5% (178) | 5.5% (91) | 0% (34) | 5.3% (38) |
|  |  | 50-80 | 5.9% (188) | 10.6% (104) | 23.1% (91) | 9.1% (77) | 14.8% (54) | 8% (25) | 20.3% (59) | 22.7% (66) | 30.8% (26) | 8.4% (381) | 9.5% (168) | 11% (154) | 3.4% (29) | 20% (20) | 26.3% (19) |
|  |  | >80th | 12.8% (78) | 27% (100) | 43.6% (117) | 12.1% (33) | 32.4% (37) | 29.4% (51) | 18.5% (27) | 29.3% (41) | 71.4% (49) | 20.2% (267) | 19% (126) | 24% (171) | 23.1% (13) | 38.5% (13) | 43.5% (23) |
| Day 4 | Position percentile | <50th | 1.6% (378) | 2.8% (216) | 14.3% (70) | 5% (101) | 14.8% (61) | 19.4% (31) | 7.7% (130) | 11.9% (67) | 17% (53) | 5.8% (521) | 8.4% (250) | 6.2% (162) | 4.3% (92) | 6.1% (33) | 2.9% (34) |
|  |  | 50-80 | 6.3% (158) | 6.8% (118) | 29.4% (68) | 4.7% (43) | 23.3% (30) | 25% (20) | 20.4% (54) | 29.8% (47) | 34.8% (23) | 9.7% (300) | 11.8% (161) | 10.8% (120) | 16.7% (24) | 12.5% (16) | 17.6% (17) |
|  |  | >80th | 22% (59) | 22.6% (62) | 52.8% (127) | 15.8% (19) | 50% (32) | 50% (30) | 34.6% (26) | 47.1% (34) | 71.1% (38) | 17.7% (175) | 24.7% (182) | 28.1% (128) | 28.6% (14) | 40% (10) | 40% (10) |
| Day 5 | Position percentile | <50th | 3.3% (273) | 7% (129) | 11.6% (43) | 8.3% (72) | 31.3% (32) | 40% (10) | 12.5% (96) | 19.1% (47) | 18.8% (32) | 6.9% (348) | 7% (273) | 10.3% (155) | 3% (67) | 4.3% (47) | 0% (15) |
|  |  | 50-80 | 9.8% (123) | 8.8% (80) | 27.1% (59) | 18.9% (37) | 26.3% (19) | 56.3% (16) | 30.8% (52) | 37% (46) | 21.4% (28) | 13.6% (221) | 8.2% (195) | 8.8% (113) | 20% (20) | 0% (16) | 35.3% (17) |
|  |  | >80th | 34.9% (43) | 31.8% (44) | 62.1% (103) | 41.2% (17) | 58.3% (12) | 40.9% (22) | 66.7% (21) | 53.3% (30) | 80% (15) | 23% (200) | 27.4% (95) | 35.2% (91) | 26.7% (15) | 20% (15) | 44.4% (9) |
| Day 6 | Position percentile | <50th | 5.1% (197) | 7.9% (76) | 13.6% (44) | 29.7% (37) | 28% (25) | 44.4% (9) | 21.3% (61) | 21.6% (37) | 15.9% (44) | 8.7% (343) | 8% (238) | 6.3% (112) | 1.5% (66) | 0% (16) | 9.5% (21) |
|  |  | 50-80 | 14.9% (87) | 25.4% (59) | 37.5% (48) | 32.3% (31) | 56.3% (16) | 38.5% (13) | 33.3% (39) | 50% (20) | 39.3% (28) | 11.5% (218) | 13.6% (154) | 11.1% (72) | 4.2% (24) | 17.6% (17) | 6.7% (15) |
|  |  | >80th | 44.1% (34) | 59.2% (49) | 67.9% (56) | 71.4% (14) | 50% (6) | 33.3% (6) | 76.9% (26) | 82.4% (17) | 62.5% (16) | 27.2% (180) | 24.6% (65) | 25.8% (66) | 12.5% (8) | 57.1% (14) | 50% (10) |

**eTable 5 – Adverse outcomes likelihood stratified by distance and position percentiles for various inflammatory cohort.**

**1.14 Routine measurement units and reference intervals**

Results in this study were generated through analysis of 20 different markers derived from the complete blood count and basic metabolic panel. In **eTable 6**, we give a brief overview of each marker, along with its standard units, and reference interval at both Massachusetts General Hospital (MGH) and Brigham and Women’s Hospital (BWH).

**eTable 6 – Routine measurement units and reference intervals**

|  |  |  | Reference interval | | | |
| --- | --- | --- | --- | --- | --- | --- |
|  |  |  | MGH | | BWH | |
|  | Abbreviation | Units | Male | Female | Male | Female |
| **Complete blood count** |  |  |  |  |  |  |
| Hematocrit | HCT | % | 41-53 | 36-46 | 40-54 | 36-48 |
| Hemoglobin | HGB | g/dL | 13.5-17.5 | 12.0-16.0 | 13.5-18 | 11.5-16.4 |
| Mean corpuscular hemoglobin | MCH | pg | 26-34 | 26-34 | 27-32 | 27-32 |
| Mean corpuscular hemoglobin concentration | MCHC | g/dL | 31-37 | 31-37 | 32-36 | 32-36 |
| Mean corpuscular volume | MCV | fL | 80-100 | 80-100 | 80-95 | 80-95 |
| Mean platelet volume | MPV | fL | 8.4-12.0 | 8.4-12.0 | 8.4-12.0 | 8.4-12.0 |
| Platelet count | PLT | 10^3^/µL | 150-400 | 150-400 | 150-400 | 150-400 |
| Red blood cell count | RBC | 10^6^/µL | 4.5-5.9 | 4.0-5.2 | 3.9-6.0 | 4.5-6.4 |
| Red cell distribution width | RDW | % | 11.5-14.5 | 11.5-14.5 | 11.5-14.5 | 11.5-14.5 |
| White blood cell count | WBC | 10^3^/µL | 4.5-11.0 | 4.5-11.0 | 4.0-10.0 | 4.0-10.0 |
| **Basic metabolic panel** |  |  |  |  |  |  |
| Anion gap | ANION | mmol/L | 3-17 | 3-17 | 3-17 | 3-17 |
| Blood-urea nitrogen | BUN | mg/dL | 8-25 | 8-25 | 6-23 | 6-23 |
| Calcium | CA | g/dL | 8.5-10.5 | 8.5-10.5 | 8.8-10.7 | 8.8-10.7 |
| Chloride | CL | mmol/L | 98-108 | 98-108 | 98-107 | 98-107 |
| Carbon dioxide | CO2 | mmol/L | 23-32 | 23-32 | 22-31 | 22-31 |
| Creatinine | CRE | mg/dL | 0.6-1.5 | 0.6-1.5 | 0.5-1.2 | 0.5-1.2 |
| Estimated glomerular filtration rate | eGFR | mL/min/1.73m^2^ | >60 | >60 | >60 | >60 |
| Glucose | GLU | mg/dL | 70-110 | 70-110 | 70-110 | 70-110 |
| Potassium | K | mmol/L | 3.4-5.0 | 3.4-5.0 | 3.4-5.0 | 3.4-5.0 |
| Sodium | NA | mmol/L | 135-145 | 135-145 | 136-145 | 136-145 |

**1.15 Validation with non-interpolated laboratory values**

Results in **Fig. 3** of the main manuscript were calculated using patient WBC and PLT values which had been interpolated and evenly sampled every 12hrs (over the first 20 days of hospital stay). This interpolation was linear, meaning interpolated values at a given timepoint rely on a blood count measurement from before and after the interpolation time. (i.e. calculating the WBC value at day 4.5 requires knowing the last WBC measurement from before day 4.5, as well as the next measurement after day 4.5). While most patient blood counts are taken daily (or more frequently), data leakage is possible, where the interpolated values contain some information that would not necessarily be available clinically.

To account for this possibility, we repeated risk prediction for the cardiac surgery cohort using raw lab data, without any interpolation. For example, we calculated a patient’s position percentile (relative to the mean favourable trajectory) on day 4 by using the most recent available blood count that is strictly prior to day 4 (i.e. less than 96hrs post-surgery). For direction percentiles at a given time point we take the last two distinct blood counts that are strictly prior to the current time.

In **eTables 7-8** we present recreations of the adverse outcome stratifications in the cardiac surgery cohort from **Fig. 1D** and **Fig. 3B** using non-interpolated laboratory values. The use of non-interpolated values does appear to lead to a slight reduction in risk stratification. However, the degree of risk stratification using non-interpolated values is consistent with the results in the main manuscript (**Fig. 1D**, and **Fig. 3B**).

**eTable 7 – Mortality risk for cardiac surgery cohort stratified direction percentiles, using non-interpolated laboratory values**

|  | With interpolation | | | | Without interpolation | | | |
| --- | --- | --- | --- | --- | --- | --- | --- | --- |
|  | Day 3 direction percentile | | | | Day 3 direction percentile | | | |
|  | <50th | 50-80 | 80-90 | >90th | <50th | 50-80 | 80-90 | >90th |
| Mortality - % | 0.60% | 1.40% | 4% | 10% | 1.20% | 2.80% | 3.00% | 6.70% |
|  | With interpolation | | | | Without interpolation | | | |
|  | Day 5 direction percentile | | | | Day 5 direction percentile | | | |
|  | <50th | 50-80 | 80-90 | >90th | <50th | 50-80 | 80-90 | >90th |
| Mortality - % | 0.80% | 1.30% | 3% | 16.40% | 0.50% | 1.60% | 4.30% | 11.50% |

**eTable 8 – Mortality or long stay risk for cardiac surgery cohort stratified by joint position and direction percentiles, using non-interpolated laboratory values**

|  |  | Day 4 - Joint position and direction stratification | | | | | |
| --- | --- | --- | --- | --- | --- | --- | --- |
|  |  | With interpolation | | | Without interpolation | | |
|  |  | Mortality or LOS > 14 - % | | | Mortality or LOS > 14 - % | | |
|  |  | Distance percentile | | | Distance percentile | | |
|  |  | <50th | 50-80 | >80th | <50th | 50-80 | >80th |
| Position percentile | <50th | 1.60% | 2.80% | 14.30% | 3.10% | 5.50% | 20% |
|  | 50-80 | 6.30% | 6.80% | 29.40% | 8.30% | 10% | 28% |
|  | >80th | 22% | 22.60% | 52.80% | 18.50% | 20% | 47.30% |
